# Optimizing and Evaluating Nanopore-Based Targeted and Metagenomic Sequencing Workflows for Rapid Diagnosis of Acute Invasive Infections from Normally Sterile Body Fluids

**DOI:** 10.1101/2025.06.02.25328768

**Authors:** Hiu-Yin Lao, Tin-Nok Hung, Timothy Ting-Leung Ng, Wing-Yin Tam, Kam-Tong Yip, Chi-Ka Cheng, Miranda Chong-Yee Yau, Alex Yat-Man Ho, Tak-Lun Que, Kitty Sau-Chun Fung, Sandy Ka-Yee Chau, Jimmy Yiu-Wing Lam, Kristine Shik Luk, Gilman Kit-Hang Siu

## Abstract

Rapid and accurate pathogen identification is critical for managing acute invasive infections. Conventional culture methods are time-consuming, delaying effective treatment. Nanopore sequencing offers real-time, long-read capabilities suitable for clinical diagnostics, yet standardized workflows remain lacking. This study developed and evaluated two optimized nanopore sequencing workflows: Nanopore Targeted Sequencing (NTS) and Nanopore Metagenomic Sequencing (NMgS), for pathogen and antimicrobial resistance (AMR) detection in 177 normally sterile body fluid samples. NTS used multiplex PCR to amplify 16S rRNA, ITS, and 21 AMR genes, while NMgS applied host DNA depletion followed by unbiased sequencing. Both workflows were benchmarked against culture-based diagnostics.

Among the 304 species cultured from 177 body fluid samples, NTS identified 78.95%, with 77.30% meeting the threshold of relative abundance (T_RA_) of 0.058 and 71.38% having at least 10 classified reads. In comparison, NMgS identified 39.47% of cultured species at the species level and 9.54% at the genus level. Of the 28 samples containing AMR ESKAPE pathogens, NTS detected associated AMR genes in 24 samples (85.71%), while NMgS identified AMR genes linked to 9 of the 32 ESKAPE pathogens (28.13%). The turnaround times for NTS and NMgS workflows were 10.75 and 12.82 hours, respectively.

In conclusion, this study demonstrated the clinical utility of Nanopore sequencing for rapid diagnosis in clinical microbiology. The heightened sensitivity of Nanopore targeted sequencing renders it ideal for routine clinical microbiology diagnoses, whereas unbiased Nanopore metagenomic sequencing is advantageous in identifying infections of unknown etiology.

## Introduction

Conventional culture remains the cornerstone of pathogen identification in clinical microbiology laboratories. However, its prolonged incubation period significantly lengthens the sample-to-report time, which is critical in acute invasive infections where mortality rates increase with delays in effective treatments [1–3]. In contrast, sequencing-based diagnostic approaches, which bypass the need for incubation, have emerged as promising alternatives. Nanopore sequencing, in particular, offers several advantages for rapid diagnosis, including long-read capabilities, portability, scalability, flexible throughput, and real-time data acquisition. Its clinical utility for rapid pathogen detection has been demonstrated in multiple pilot studies employing either targeted [4–7] or metagenomic sequencing approaches [8–10]. Nevertheless, the lack of standardized protocols and regulatory guidelines continues to hinder the routine implementation of sequencing-based diagnostics in clinical microbiology practice.

To address these challenges, optimized workflows for both nanopore targeted sequencing (NTS) and nanopore metagenomic sequencing (NMgS) were developed in this study, encompassing steps from DNA extraction to data analysis. In NTS workflow, a set of identification (ID) primers was used to amplify 16S rRNA gene and internal transcribed spacer (ITS) regions for bacterial and fungal identification, respectively. Additionally, a panel of antimicrobial resistance (AMR) primers was employed to detect 19 clinically important AMR genes commonly associated with ESKAPE pathogens, particularly those mediating plasmid-borne resistance to β-lactams and vancomycin. For the NMgS workflow, host DNA depletion was performed prior to DNA extraction using 0.0125% of saponin [11] for differential lysis of human cells, followed by treatment with PMAxx™ dye to crosslink and inactivate DNA from non-viable cells [12], thereby preventing its amplification and sequencing.

NTS selectively amplifies multiple targeted sequences, enhancing sensitivity and reducing sequencing depth requirements, though it is limited to organisms and genes included in the panel [13]. In contrast, NMgS enables comprehensive and unbiased profiling by sequencing all genetic material in a sample, but its sensitivity is often compromised by the high proportion of host DNA in clinical specimens. To evaluate the clinical utility of both workflows for pathogen identification and antimicrobial resistance detection, a total of 177 normally sterile body fluids were collected from 4 public hospitals in Hong Kong and sequenced using both NTS and NMgS workflows. Sequencing was performed for four hours in both workflows to simulate a rapid diagnostic setting, and results were compared with those from routine culture. Additionally, 69 samples underwent extended NMgS sequencing for up to 48 hours to evaluate the impact of sequencing duration on sensitivity, with results at 24 and 48 hours compared against culture-based findings.

## Materials and methods

### Sample collection

A total of 177 residual normally sterile body fluids with positive culture results were collected from 4 public hospitals, including Tuen Mun Hospital, United Christian Hospital, Pamela Youde Nethersole Eastern Hospital, and Princess Margaret Hospital. Each sample was aliquoted into two portions, with one processed using the NTS workflow and the other using the NMgS workflow.

### Nanopore targeted sequencing (NTS) workflow

A detailed protocol for NTS workflow is provided in Supplemental 1. DNA was extracted using the QIAamp BiOstic Bacteremia DNA Kit. For each sample, two multiplex PCR reactions were prepared: ID PCR and AMR PCR. While the ID PCR amplified 16S rRNA gene and ITS gene, the AMR PCR amplified 19 clinically important beta-lactam resistance genes (*MecA, blaCTX-M, blaTEM, blaSHV, blaOXA-1, blaKPC, blaIMP, blaVIM, blaNDM, blaOXA-48, blaACC, blaFOX, blaMOX, blaCMY, blaDHA, blaLAT, blaBIL, blaMIR,* and *blaACT*) and 2 vancomycin resistance genes (*VanA* and *VanB*). PCR products of each sample were pooled and purified using 0.7x AMPure XP beads.

Library preparation was conducted using the ligation sequencing kit (SQK-LSK110 for R9.4.1 or SQK-LSK114 for R10.4.1 flow cells) and PCR Barcoding Expansion 1–96 kit (EXP-PBC096), with minor modifications. Briefly, barcoding PCR was performed, followed by purification with 0.8x AMPure XP beads. After quantification and normalization, up to 24 samples were pooled, end-repaired, adapter-ligated, and sequenced on a GridION for 4 hours using super accuracy basecalling (SUP) model.

Sequencing reads were analyzed using an in-house pipeline (https://github.com/siupenyau/idamr). Reads were first filtered by length using NanoFilt v2.8.0 [14] , separating them into two groups: 1300–1700 bp (16S rRNA genes) and 100–1200 bp (ITS and AMR genes). Taxonomic classification of 16S rRNA genes was performed using Emu v3.4.5 [15]. ITS and AMR reads were classified using BLAST+ [16] with the ITS database built based on NCBI Fungal ITS RefSeq Targeted Loci Project (PRJNA177353) and the NCBI AMR database Reference Gene Catalog [17], respectively. BLASTn searches were conducted with an e-value threshold of 1e−5 and a minimum identity of 90% [18].

### Validation of NTS workflow

The NTS workflow was validated using simulated bacteremic and fungemic blood samples. Four microorganisms with distinct cell wall structures — *Staphylococcus aureus* ATCC BAA-3114, *Escherichia coli* ATCC BAA-3054, *Mycobacterium marinum* ATCC BAA-535, and *Candida krusei* ATCC 6258, were individually spiked into whole blood at a final concentration of 100 CFU/mL. The NTS workflow was then applied to detect the presence of each organism, using a previously established threshold of relative abundance (T_RA_ = 0.058) for species identification [19]. Additionally, AMR gene detection was performed for the antimicrobial-resistant strains (*S. aureus* and *E. coli*).

### Nanopore metagenomic sequencing (NMgS) workflow

A detailed protocol for NMgS workflow is provided in Supplemental 2. Host DNA depletion was conducted before DNA extraction. For blood samples, additional hetasep treatment was performed to remove red blood cells prior to depletion. Body fluids were initially centrifuged to collect the pellets, followed by osmotic lysis using 1 ml of nuclease-free water. The samples were then centrifuged again, the resulting pellets were resuspended in 0.0125% saponin [11] and then treated with 20 μM PMAxx [12]. After that, samples were washed with 1 ml saline and subjected to DNA extraction using the QIAamp BiOstic Bacteremia DNA Kit.

Library preparation for NMgS workflow was conducted using the ligation sequencing kit (SQK-LSK110 or SQK-LSK114) and PCR Barcoding Expansion 1-96 (EXP-PBC096), with modifications. Genomic DNA was fragmented and A-tailed using the NEBNext Ultra II FS DNA module, followed by purification with 1x AMPure XP beads. Barcode adapter ligation was performed, followed by barcode PCR, and amplicons were purified using 0.8x AMPure XP beads. After quantification and normalization, the pooled library was end-repaired, adapter-ligated, and sequenced on GridION using SUP model for 4 hours or up to 48 hours. Adaptive sampling was enabled to deplete reads mapped to human reference genome GRCh38.p13.

Sequencing reads were analyzed using an in-house pipeline (https://github.com/siupenyau/idamr_meta). Reads shorter than 200 bp were removed, followed by human read removal using Kraken 2 v2.1.3 [20] with the human genome database. The remaining reads were classified using Kraken 2 with the PlusPF database, and species abundance was re-estimated using Bracken v2.9 [21]. For AMR gene identification, filtered reads were classified using BLAST+ [16] with the NCBI Reference Gene Catalog [17] as in the NTS workflow.

### Validation of NMgS workflow

The NMgS workflow was validated using simulated bacteremic and fungemic blood samples as in the NTS workflow. However, the final concentration of spiked microorganisms was adjusted to 1000 CFU/ml. To assess the effect of host DNA depletion, each spiked sample was divided into two groups: one subjected to human DNA depletion and the other left untreated. After library preparation, the pooled library was sequenced for 4 hours using SUP basecalling mode with adaptive sampling. The relative abundance of human DNA in treated and control samples was compared.

To evaluate the optimal confidence threshold of Kraken 2 for classifying low microbial biomass clinical specimens, the reads were analyzed across a range of confidence thresholds from 0 to 1, with intervals of 0.05. Additionally, the impact of the read threshold in Bracken was evaluated at values of 0, 5, and 10. The optimal threshold for both Kraken 2 and Bracken was defined as the minimum value that yielded the highest abundance of spiked organisms and the lowest background noise for the four spiked species.

### Clinical Utility Evaluation of NTS and NMgS Workflows

To assess the clinical utility of NTS and NMgS workflows, sequencing was performed on 177 sterile body fluid samples using both methods. To ensure timely results, all samples underwent a standard sequencing duration of 4 hours. Additionally, 69 samples in the NMgS workflow were subjected to extended sequencing for up to 48 hours to evaluate potential improvements in sensitivity. Sequencing data were analyzed using an in-house bioinformatics pipeline, and results were compared with reference culture findings to determine diagnostic accuracy.

## Results

### Validation of NTS workflow

To validate the NTS workflow, four microorganisms with distinct cell wall structures were individually spiked into blood samples in triplicate. Their average relative abundances were then determined. A minimum threshold of 0.058 for relative abundance was applied [19]. All spiked organisms were successfully detected using the NTS workflow. The average relative abundance ± standard deviation (SD) for *S. aureus*, *E. coli*, *M. marinum*, and *C. krusei* was 0.495 ± 0.153, 0.305 ± 0.142, 0.174 ± 0.141, and 0.729 ± 0.114, respectively.

For the antimicrobial resistant ATCC reference strains *S. aureus* (BAA-3114) and *E. coli* (BAA-3054), all targeted antimicrobial resistance (AMR) genes were detected in the NTS workflow with more than 10 reads. The mean read count ± SD for the mecA gene in *S. aureus* was 18 ± 2.94. For *E. coli*, the mean read counts ± SD for the CTX-M and OXA-1 genes were 51.67 ± 30.92 and 12.33 ± 0.94, respectively.

### Validation and optimization of NMgS workflow

Similar to the NTS workflow, NMgS workflow was also validated using blood samples spiked with *S. aureus*, *E. coli*, *M. marinum*, and *C. krusei*. For each microbial suspension, two spiked blood samples were prepared: one went for the saponin-based host DNA depletion, while the other served as untreated control.

After 4 hours of sequencing, the untreated control samples yielded an average of 9036.58 reads, with a SD of 5132.72 and a median of 8399.50. In contrast, the host DNA-depleted samples had a mean total read count of 5,246.67 ± 2,076.02, with a median of 4348. After filtering out reads below 200 bp, the mean number of reads ± SD for control and treated samples were 8548.17 ± 4952.59 and 3816.33 ± 1807.33, respectively. The corresponding medians were 7,993.50 and 3,059.50.

Figure 1 illustrates the relative abundance of human DNA in control and host DNA-depleted samples. On average, the percentage of human reads decreased from 99.5% to 49.1%, highlighting the effectiveness of the saponin-PMA-based host DNA depletion method. The most significant reduction was observed in samples spiked with *M. marinum* (reduced from 99.6% to 24.0%), followed by *E. coli* (reduced from 99.4% to 43.5%). Reductions in samples spiked with *C. krusei* and *S. aureus* were similar, decreasing from 99.6% to 63.7% and from 99.5% to 62.3%, respectively.

**Figure 1:**
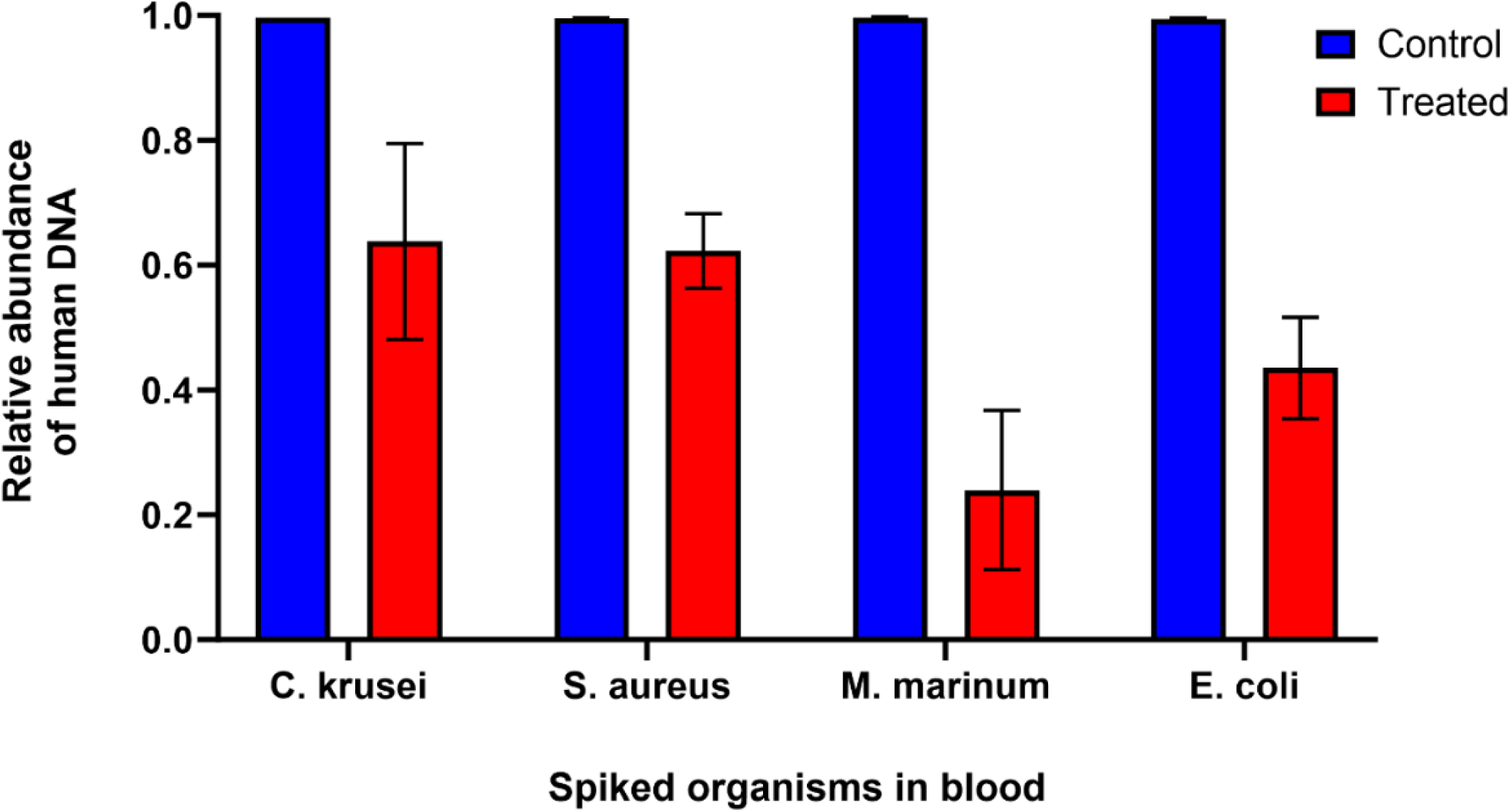
The relative abundance of human DNA in control and treated samples.

Following the removal of human reads, Kraken 2 was used for taxonomic classification. To determine the optimal confidence threshold of Kraken 2 for classifying low microbial biomass clinical samples, thresholds ranging from 0 to 1 (in 0.05 increments) were evaluated. Additionally, the impact of read number thresholds in Bracken was assessed at values of 0, 5, and 10. While increasing these thresholds reduced false positives, it also decreased the number of classified reads and therefore the sensitivity. Based on the analysis of relative abundance changes across different thresholds (Figure 2), the optimal settings were determined to be a confidence threshold of 0.1 for Kraken 2 and a read count threshold of 10 for Bracken. These settings were subsequently applied in the NMgS workflow for downstream analysis.

**Figure 2:**
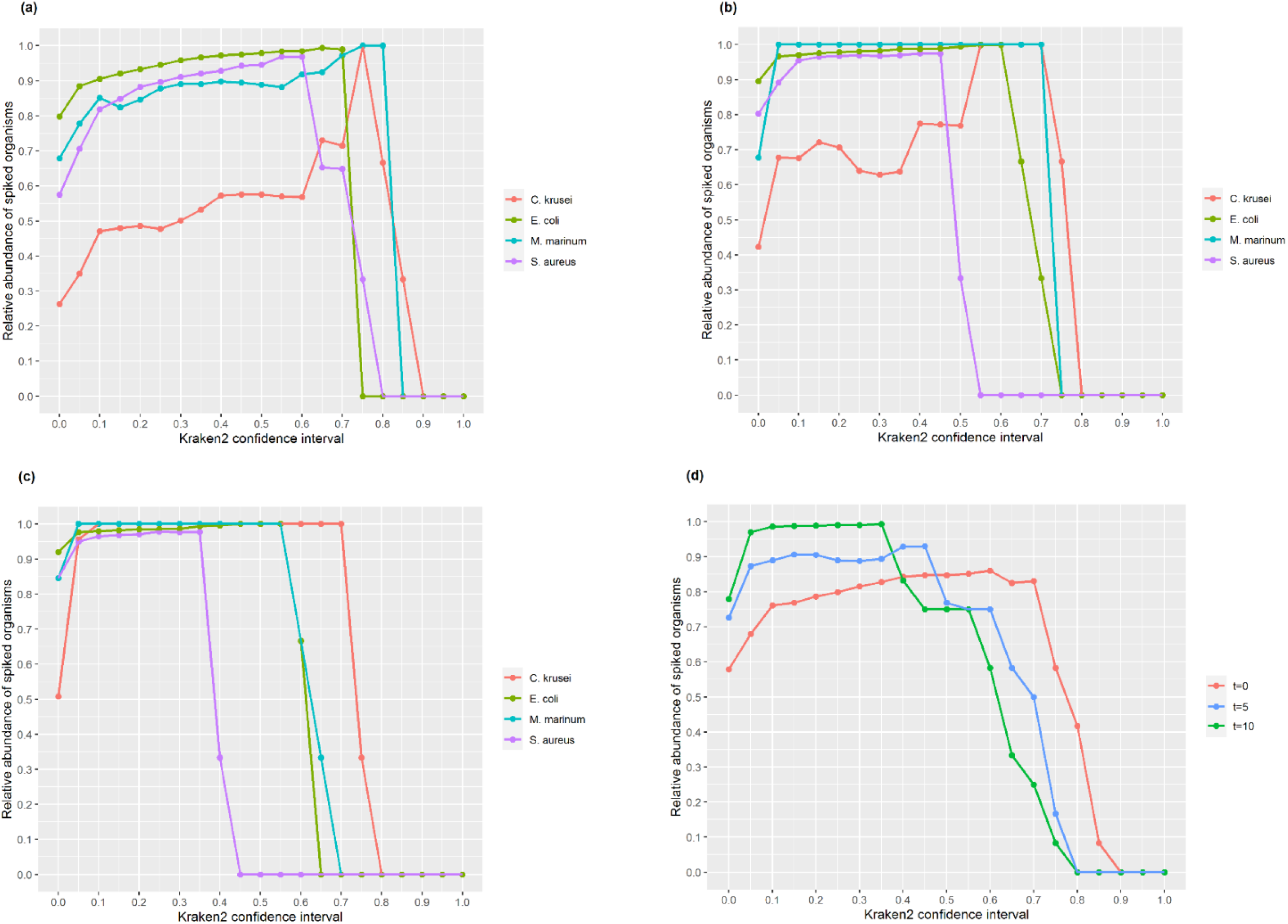
The change in relative abundance of each spiked organism across a range of confidence thresholds of Kraken 2 and threshold of Bracken at (a) t =0, (b) t = 5, and (c) t = 10; (d) shows the average relative abundance of the four organisms varies against confidence thresholds of Kraken 2 and threshold of Bracken.

Figure 3 compares the relative abundance of spiked organisms in control and host DNA-depleted samples, with and without applying the optimal thresholds. In control samples, the average count of non-human reads was notably low, at 37.25 ± 18.78, with a median of 30. In contrast, the host DNA-depleted samples showed a significantly higher average of 1,899.75 non-human reads, with a SD of 1051.44 and a median of 1694. After applying the optimal thresholds, the relative abundance of spiked organisms increased from an average of 57.9% to 98.6%.

**Figure 3:**
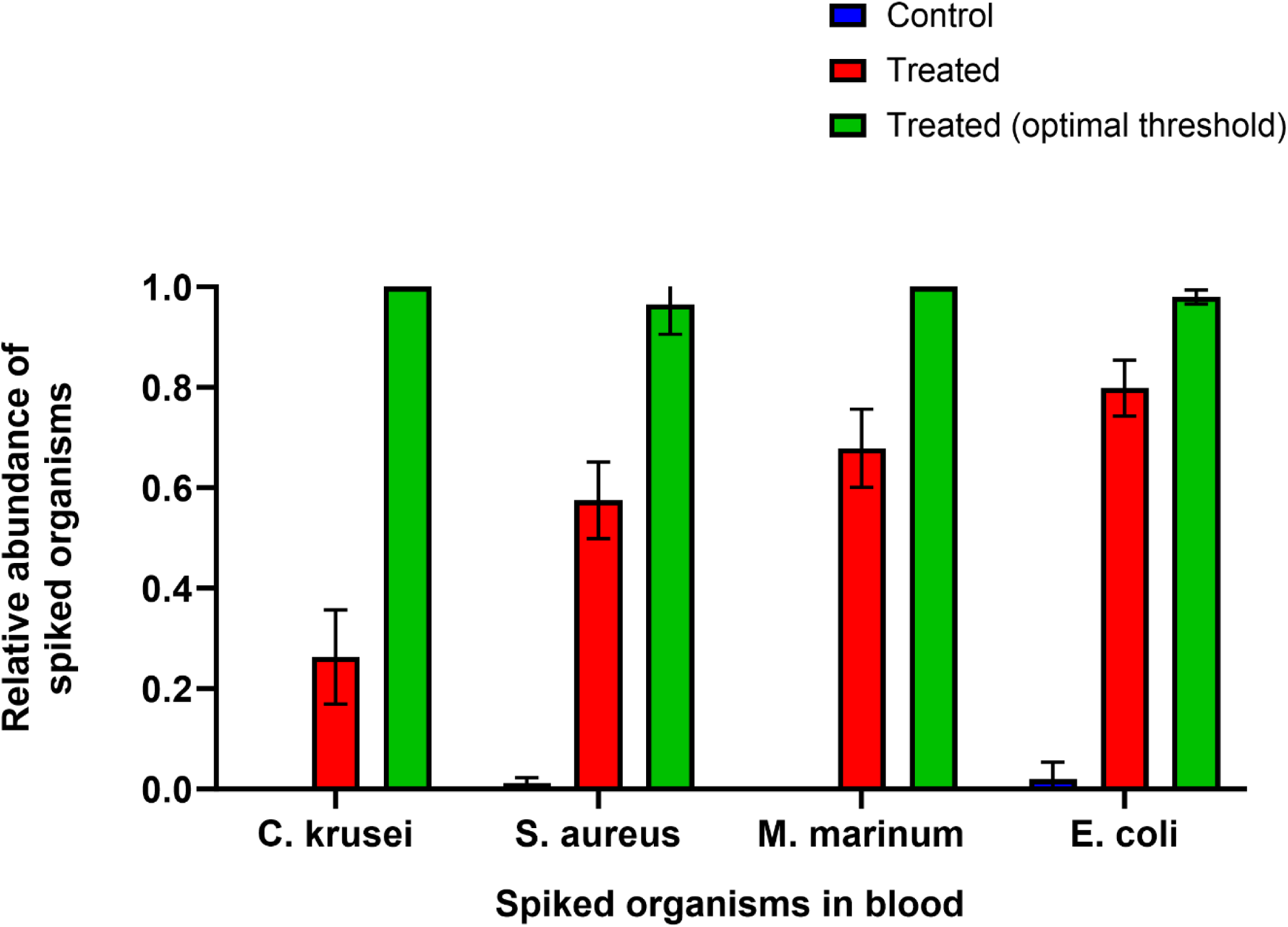
The relative abundance of spiked organisms after removal of human reads.

### The concordance between NTS and culture in pathogen identification

A total of 177 samples were sequenced using NTS workflow, including 119 monomicrobial samples and 58 polymicrobial samples. The concordance between NTS and culture for pathogen identification in each samples is summarized in Supplemental 3. Figure 4a and 4b illustrate the overall concordance between NTS and culture for pathogen identification in monomicrobial and polymicrobial samples, respectively.

**Figure 4:**
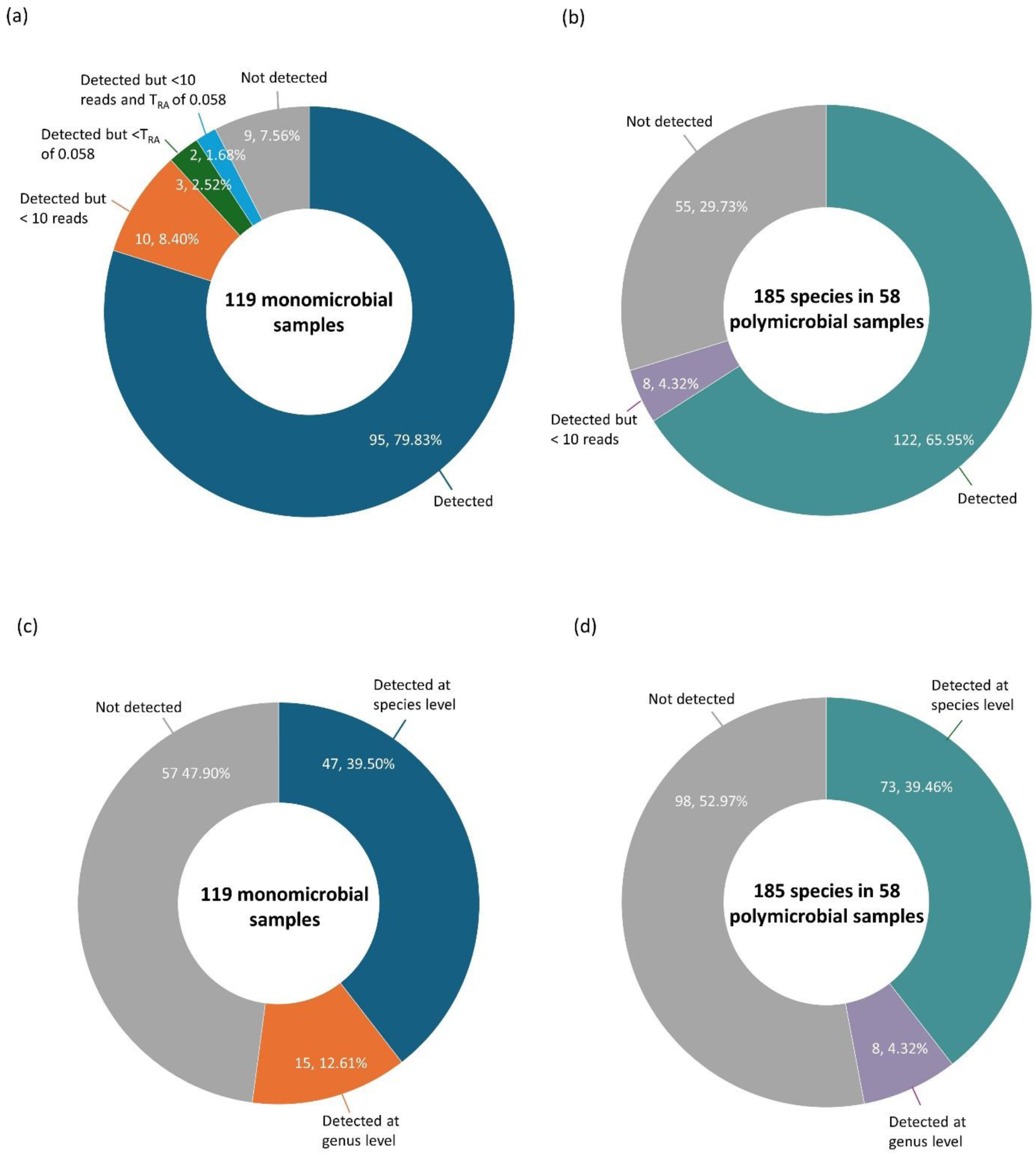
Concordance between NTS and culture for pathogen identification in (a) monomicrobial and (b) polymicrobial samples, and between NMgS and culture in (c) monomicrobial and (d) polymicrobial samples.

Among the 119 monomicrobial samples, 116 were identified as bacterial infections and 3 as fungal infections by culture. NTS detected at least one read corresponding to the culture-identified species in 110 samples (92.44%). When applying a T_RA_ of 0.058, the targeted species were detected in 105 samples (88.24%). Of the 110 concordant samples, 12 exhibited very low read counts (<10 reads) for the targeted species, with two of these falling below the T_RA_. In the 9 discordant samples (7.56%), two showed no classified reads at all. Environmental contaminants were identified in six out of nine samples, including but not limited to *Pelomonas saccharophila*, *Ralstonia insidiosa*, *Burkholderia stabilis*, and *Sphingomonas lacus*. In the remaining one discordant sample, 15 unexpected species were detected, predominantly anaerobes such as *Bacteroides fragilis*, *Peptostreptococcus stomatis*, *Prevotella nigrescens*, and *Faecalibacterium prausnitzii*.

In monomicrobial samples, the mean number of classified reads for the targeted species was 4094.41 ± 7271.79, with a median of 824. The mean number of classified bacterial species identified by Emu was 4.20 ± 7.51, with a median of 2. For ITS analysis, there were 44 samples (36.97%) yielded at least one classified read. Among these, 30 samples (68.18%) exhibited very low total classified reads (<10 reads), primarily consisting of background contaminants such as *Cladosporium antarcticum* and *Aspergillus versicolor*. Of the three culture-confirmed fungal infections, NTS detected the corresponding fungal species in two samples, although one had fewer than 10 classified reads. In non-fungal infection samples with more than 10 classified reads, opportunistic fungal pathogens such as *Trichosporon asahii* and *Malassezia arunalokei* were identified alongside environmental contaminants.

Among the 58 polymicrobial samples, 48 were bacterial infections, 2 involved dual fungal infections, and 8 were bacterial and fungal co-infections. NTS detected all targeted species in 23 samples (39.66%), and at least one targeted species in the remaining 35 samples (60.34%). Culture identified a total of 185 species across these samples, of which NTS detected 130 (70.27%). Eight of the detected species had very low read counts (<10 reads). The mean number of classified reads for targeted species was 2,293.90 ± 5,822.54, with a median of 228. Emu classified an average of 18.64 bacterial species per sample (SD: 23.22; median: 11.50). ITS analysis yielded at least one classified read in 16 samples (27.59%), with 6 of these having fewer than 10 reads. NTS detected 10 out of 12 fungal species identified by culture, with two of them having classified reads below 10.

Across 177 body fluid samples, the overall concordance between NTS and culture for pathogen identification was 78.95% (240/304) when detection was defined as at least one read of the targeted species. Applying a T_RA_ of 0.058 to monomicrobial samples reduced the concordance to 77.30% (235/304). When an additional threshold of ≥10 reads was applied, the concordance further decreased to 71.38% (217/304).

### The concordance between NMgS and culture in pathogen identification

The concordance between NMgS and culture for pathogen identification in each sample was detailed in Supplemental 4. An overview of the overall concordance between NMgS and culture for pathogen identification in both monomicrobial and polymicrobial samples is illustrated in Figure 4c and 4d, respectively.

For 119 monomicrobial samples, NMgS correctly detected 47 species (39.50%) and classified an additional 15 samples (12.61%) at the genus level after 4 hours of sequencing. The mean read count for the targeted species was 9060.51, with a SD of 18162.25 and a median of 705. Notably, 56 samples (47.06%) yielded no classified reads after applying the optimal thresholds. Among the remaining 63 samples, the mean number of classified species was 2.23, with a SD of 2.04 and a median of 1.

In the 58 polymicrobial samples, NMgS successfully identified all targeted species or their genus in 12 samples (20.69%) and at least one but not all in 28 samples (48.28%). Of the 18 samples (31.03%) with no matches at all, 12 samples comprising 48 cultured species had no classified reads after four hours of sequencing. Among the 46 samples with at least one match, the average number of classified species was 8.21, with a standard deviation of 9.11 and a median of 5. Out of the 185 species cultured from the 58 polymicrobial samples, NMgS correctly detected 73 species (39.46%) and identified an additional 14 cultured species (7.57%) at the genus level only.

Overall, NMgS correctly detected 120 species (39.47%) out of 304 species cultured across all 177 samples and classified an additional 29 cultured species (9.54%) at the genus level. A total of 155 species (50.99%) were not detected by NMgS workflow after 4 hours of sequencing.

To assess the impact of extended sequencing duration on the sensitivity of the NMgS workflow, a subset of 69 samples was sequenced for up to 48 hours. Among the 115 cultured species from these samples, NMgS detected 55 species (47.83%) or their genus after 4 hours. After 24 hours, an additional 14 species were identified, increasing the cumulative detection rate to 60%. By 48 hours, 8 more species were detected, bringing the total to 66.96%. These findings suggest that prolonged sequencing enhances the sensitivity of the NMgS workflow.

### The concordance between NTS, NMgS and culture in AMR ESKAPE pathogens identification

A total of 32 antimicrobial-resistant ESKAPE pathogens were identified in 28 out of 177 samples using culture-based methods. These included 14 ESBL-producing *Enterobacterales*, 14 methicillin-resistant *Staphylococcus aureus* (MRSA), and 4 carbapenem-resistant organisms. No vancomycin-resistant Enterococcus (VRE) was detected in this study. Overall, NTS detected AMR genes in 24 out of 28 samples (85.71%), whereas NMgS identified 9 out of the 32 ESKAPE pathogens (28.13%). The corresponding AMR genes detected by NTS and NMgS are detailed in Table 1.

**Table 1:**
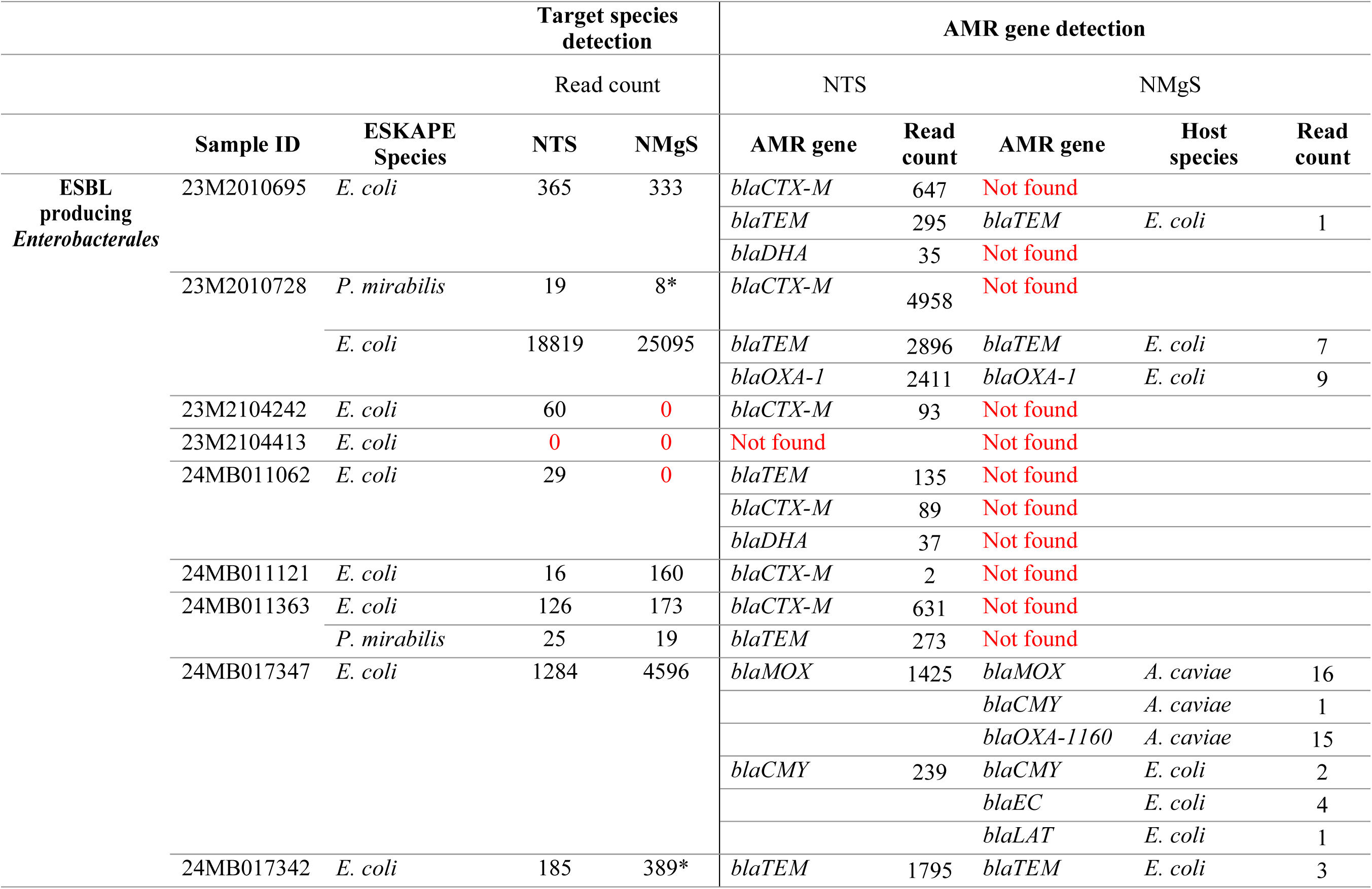
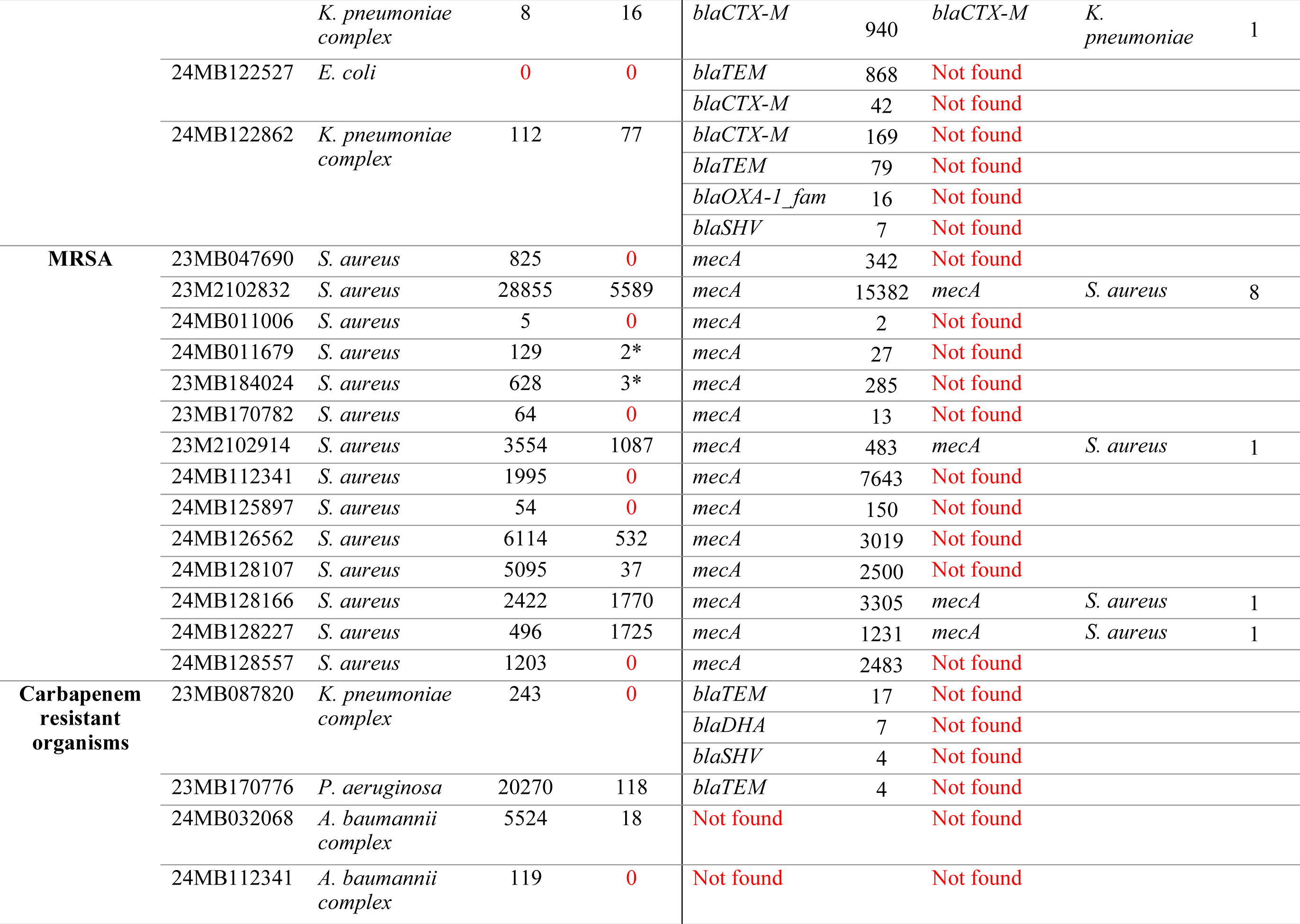
The AMR genes detected by NTS and NMgS in 28 samples with ESKAPE pathogens. *Detected only when no threshold was applied in Bracken.

### Detection of ESBL and *AmpC* genes

Among the 14 ESBL-PE strains identified in 11 samples via culture, NTS detected 12 (85.71%) of the targeted species and identified ESBL genes in 10 samples (90.91%). Notably, one sample contained only AMR genes without detection of the corresponding targeted species. The most frequently detected ESBL gene was *blaCTX-M* (9 samples), followed by *blaTEM* (7 samples).

In contrast, the NMgS workflow identified 10 targeted species but detected only 5 ESBL-PE strains (35.71%) across 4 samples (36.36%). These included four *E. coli* strains and one *K. pneumoniae* strain. The most prevalent ESBL gene detected by NMgS was *blaTEM*, found in 3 samples.

In a polymicrobial sample (24MB017347) containing ESBL-producing *E. coli*, both NTS and NMgS identified *blaAmpC* genes, specifically *blaMOX* and *blaCMY.* Notably, NMgS revealed that the *blaMOX* gene originated from *Aeromonas caviae*, another pathogen present in the sample. *Aeromonas* species are known to harbor chromosomal *blaAmpC* genes and are hypothesized to be the source of CMY-1/MOX-family enzymes [22]. Moreover, NMgS identified the *blaCMY* gene in both *A. caviae* and *E. coli*, and exclusively detected *blaEC* and *blaLAT* in *E. coli*. This highlights that while NTS solely confirmed the existence of AMR genes, NMgS had the capability to trace the source of these AMR genes.

### Detection of *mecA* genes

Among the 14 MRSA strains identified by culture, NTS successfully detected both *Staphylococcus aureus* and the mecA gene in all corresponding samples (100%). However, one sample yielded fewer than 10 classified reads for both *S. aureus* and *mecA*. In contrast, NMgS identified *S. aureus* in 8 out of the 14 samples (57.14%), with two of these detections occurring only when no read threshold was applied in Bracken. The mecA gene was detected by NMgS in only 4 samples (28.57%), all of which had high *S. aureus* read counts (>1,000 reads). Nevertheless, in 3 of these 4 samples, only a single read corresponding to the mecA gene was identified.

### Detection of carbapenemase genes

A total of four carbapenem-resistant organisms were identified by culture, including one *Klebsiella pneumoniae,* one *Pseudomonas aeruginosa*, and two *Acinetobacter baumannii* strains. While all species were detected by NTS, no carbapenemase genes were identified. Instead, ESBL or *blaAmpC* genes were detected in the samples containing *K. pneumoniae* and *P. aeruginosa*. Specifically, *blaTEM, blaSHV* (<10 reads), and *blaDHA* (<10 reads) were found in the *K. pneumoniae* sample, while *blaTEM* (<10 reads) was detected in the *P. aeruginosa* sample. Using NMgS, only one *P. aeruginosa* and one *A. baumannii* were identified among the four carbapenem-resistant organisms. Notably, *A. baumannii* was detected with fewer than 10 classified reads. Consistent with NTS, no carbapenemase genes were detected in any of the samples by NMgS.

For the carbapenem-resistant *Klebsiella pneumoniae*, PCR tests were conducted by the clinical laboratory to validate the presence of carbapenemase genes, including *blaGES, blaIMI, blaKPC, blaNmcA, blaSME, blaGIM, blaIMP, blaNDM, blaSIM, blaSPM, blaVIM*, and *blaOXA-48-like*. However, none of these genes were detected, aligning with the sequencing results. This suggests that carbapenem resistance in this strain may be mediated by mechanisms other than carbapenemase production.

### Turnaround time of NTS and NMgS workflows

The workflow and required time for each step in both NTS and NMgS are outlined in Figure 5. Compared to the 24-48 hours of incubation time needed for culture, the turnaround time for both NTS and NMgS workflows was significantly shorter, at 10.75 hours and 12.82 hours, respectively. Similar to the 16S Barcoding Kit 1-24 (SQK-16S024), a batch of 24 samples were sequenced per run in NTS workflow. In contrast, NMgS workflow accommodated up to 16 samples per run to enhance sequencing depth per sample and sensitivity. In this study, both NTS and NMgS workflows were sequenced up to 4 hours in order to have rapid diagnosis. Notably, nanopore sequencing enables real-time sequencing, allowing the extraction of fastq files at any point during the process for immediate analysis. Furthermore, nanopore flow cell is reusable and can be used up to 72 hours, enabling multiple runs with a single flow cell. The analysis for both workflows could be completed within an hour in general, depending on factors such as the number of reads per sequencing run and available computing resources.

**Figure 5:**
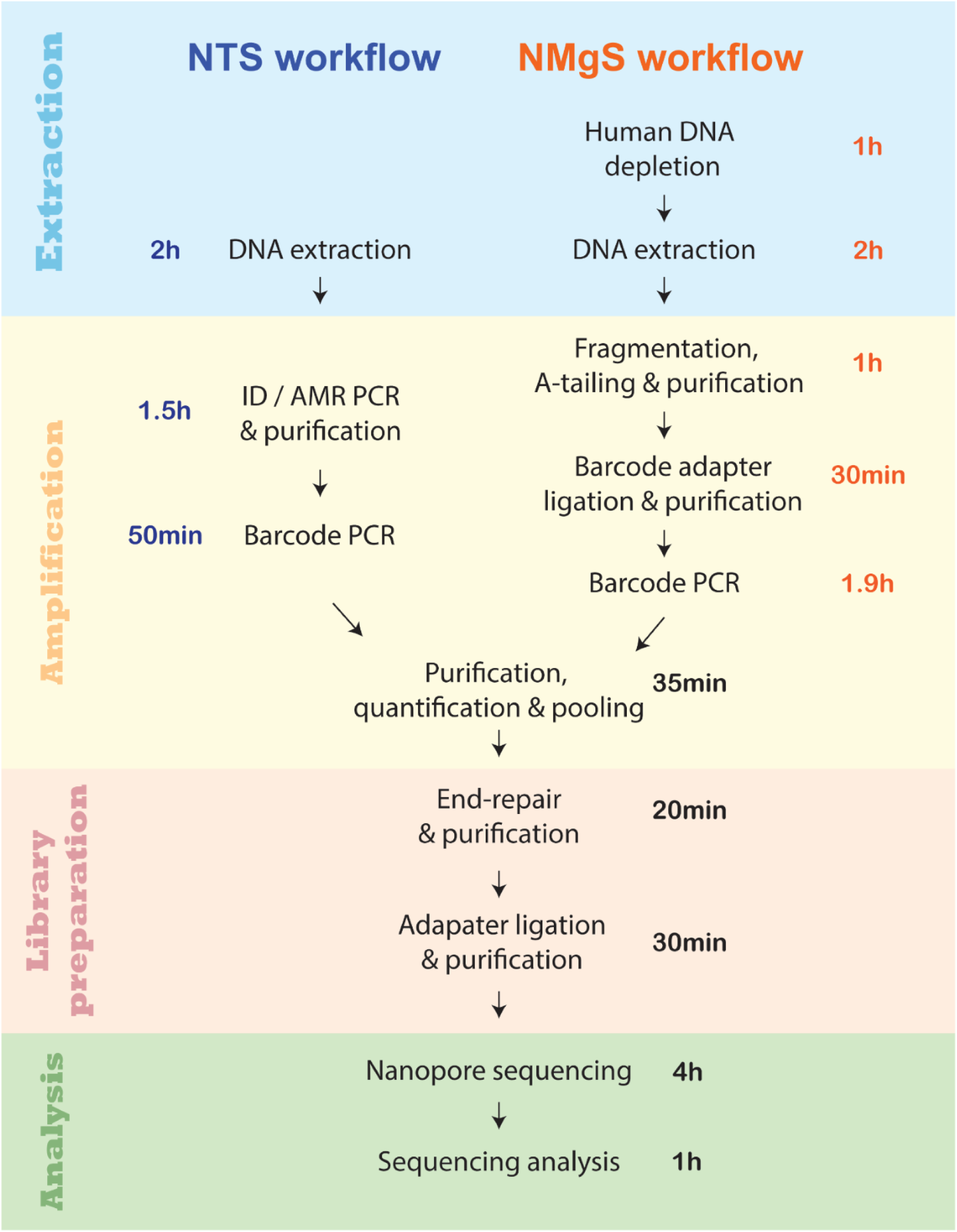
The workflow of NTS and NMgS.

## Discussion

In clinical microbiology, one of the primary challenges of sequencing-based diagnostics lies in the absence of standardized protocols and guidelines for data interpretation. This study aimed to develop an optimized protocol for nanopore sequencing-based diagnostics. Two distinct workflows were established: nanopore targeted sequencing (NTS) and unbiased nanopore metagenomic sequencing (NMgS). A pilot study was conducted to assess their performance in pathogen identification and antimicrobial resistance (AMR) detection across 177 normally sterile body fluid samples, using traditional culture-based methods as a reference. The findings offer valuable insights into the development of standardized sequencing protocols and highlight the respective advantages and limitations of targeted versus unbiased metagenomic approaches in clinical diagnostics.

The NTS workflow demonstrated high sensitivity for bacterial and fungal detection through amplification of conserved genetic markers (16S rRNA and ITS). However, its scope is inherently limited by the predefined primer panel, which excludes viral pathogens due to the absence of a universal viral barcode and the additional complexity introduced by reverse transcription for RNA viruses. While real-time PCR or immunoassays remain more practical for targeted viral detection, the lack of viral coverage in NTS restricts its utility in comprehensive infectious disease diagnostics. Furthermore, although the multiplex PCR-based AMR panel enabled rapid detection of clinically relevant resistance genes, it did not allow for the determination of the host organism, limiting its epidemiological and therapeutic value.

In contrast, the NMgS workflow offers a hypothesis-free approach capable of detecting a broad spectrum of pathogens and AMR genes. The incorporation of a saponin-PMA-based host DNA depletion step significantly improved microbial signal by reducing human DNA content. Saponin, a natural detergent, selectively disrupts the lipid bilayer of human cell membranes while exerting minimal effects on the structurally distinct bacterial and fungal cell walls. However, its working concentration varies (0.0125% - 2.2%) across studies [11, 23], and higher concentrations have been reported to compromise the integrity of gram-negative bacteria, potentially biasing microbial community profiles toward gram-positive taxa [24]. To minimize such bias, a conservative concentration of 0.0125% saponin was employed in the NMgS workflow. Following saponin treatment, propidium monoazide, a photo-reactive DNA-binding dye, was used to selectively inactivate free DNA from lysed host cells [25]. It is a more efficient and economical approach for degrading host DNA compared to DNases [26]. In the validation study of NMgS workflow, the combination of 0.0125% saponin and 20μM PMA effectively reduced human DNA in blood samples spiked with low microbial content (1000 CFU/ml) from an average of 99.5% to 49.1%.

In normally sterile body fluids, the presence of any microorganisms is generally considered indicative of infection. However, sequencing-based approaches are highly susceptible to environmental DNA contamination. Establishing a detection threshold can help differentiate true pathogens from false positives, but the optimal threshold may vary depending on the sequencing platform, analytical pipeline, and sample type. In the NTS workflow, a threshold relative abundance (T_RA_) of 0.058 [19] was applied to differentiate potential pathogens from background contaminants in monomicrobial samples. For the NMgS workflow, optimal thresholds were determined to be 0.1 for Kraken 2 and 10 classified reads for Bracken. Besides, to minimize contamination in sequencing-based diagnostics, it is essential to maintain a clean work environment, separate pre- and post-PCR areas, and include both extraction and non-template controls in the workflow.

Bioinformatic tools and reference databases significantly influence taxonomic and AMR gene classification, underscoring the need for standardized analytical pipelines in clinical settings. In the NTS workflow, read length-based segregation enabled efficient downstream analysis: reads of 100–1200 bp were used for ITS and AMR detection, while 1300–1700 bp reads were analyzed for 16S rRNA classification. Emu was selected for 16S analysis due to its high species-level resolution and concordance with culture results [19]. BLAST+, a widely recognized gold standard, was utilized for the classification of reads in ITS and AMR analysis. The overall sequencing analysis time for a batch of 24 samples was about 30 minutes to 1 hour, depending on the sequencing output.

In the NMgS workflow, adaptive sampling was employed during the sequencing to further reduce host reads, its effectiveness in depleting human reads and enriching microbial reads have been demonstrated in various studies [27–29]. Kraken 2 with Bracken was chosen for taxonomic classification in NMgS workflow due to its efficiency and computational feasibility compared to Blastn [30, 31]. Sequencing reads below 200bp and human reads were filtered out before classification. Species abundance re-estimation with Bracken was performed. This combination was found to be more accurate and less susceptible to misclassification of clinical metagenomic samples compared to using Kraken 2 alone [32]. AMR detection in NMgS also utilized BLAST+ against the NCBI Reference Gene Catalog, with host assignment inferred by reclassifying flanking sequences using the NCBI nt database. The full analysis pipeline required ∼30 minutes for 16 samples following 4 hours of sequencing.

For pathogen identification, NTS achieved an overall concordance of 78.95% (240/304) with culture, with higher agreement in monomicrobial (92.44%) than polymicrobial (70.27%) samples. NMgS showed lower concordance (67.76%), detecting 52.10% of target species in monomicrobial and 47.03% in polymicrobial samples. The reduced sensitivity of NMgS was attributed to residual host DNA masking microbial reads, particularly in low-biomass samples. Although longer sequencing runs (24–48 hours) could improve sensitivity, they are impractical for routine diagnostics.

NTS exhibited greater sensitivity than NMgS in detecting AMR ESKAPE pathogens. A key limitation of NTS is its inability to identify the specific hosts of AMR genes. Culture confirmed 12 ESBL-producing strains in 9 samples, but NTS detected ESBL genes in only 7 samples, with one below the 10-read threshold. NMgS, however, allows host classification by analyzing flanking sequences, though it identified only 4 ESBL-PE species. While a 10-read threshold improved reliability in AMR detection, single relevant reads were considered acceptable in nanopore metagenomic sequencing due to long-read lengths [33]. Thus, a sample was classified as AMR-positive with at least one relevant read. For MRSA strains, NTS showed high concordance with culture, detecting the mecA gene in 7 of 8 samples, compared to just 2 MRSA strains identified by NMgS. Regarding carbapenem-resistant organisms, neither NTS nor NMgS detected carbapenemase genes in the 4 cases. Prior PCR testing also failed to detect carbapenemase genes, suggesting resistance was acquired through other mechanisms. This highlights the challenge of inferring phenotypic resistance from genotypic data, as discrepancies between detected AMR genes and actual resistance have been reported in multiple studies [34–36]. While NMgS provides host-specific insights, NTS delivers higher sensitivity, making the balance between these factors crucial in AMR surveillance.

This study has several limitations. The optimization of NTS and NMgS workflows relied on blood samples spiked with microorganisms at known concentrations, as blood is the most accessible sterile body fluid from healthy individuals. However, no clinical blood samples were included, as standard practice involves direct inoculation into blood culture bottles to minimize contamination, leaving no residual specimens for sequencing analysis. Additionally, each spiked sample contained only a single microbial species, limiting the evaluation of workflow performance in polymicrobial infections. Moreover, the sample set was predominantly composed of bacterial infections, with limited representation of fungal pathogens. As a result, the performance of both workflows in fungal detection may not be fully characterized. Furthermore, workflow validation relied solely on comparisons with routine culture results, which often fail to detect fastidious or anaerobic organisms [37].

## Conclusion

This study developed and evaluated two nanopore-based sequencing workflows—Nanopore Targeted Sequencing (NTS) and Nanopore Metagenomic Sequencing (NMgS)—for pathogen and antimicrobial resistance (AMR) detection, using culture as the reference standard. NTS demonstrated higher sensitivity and concordance with culture due to targeted amplification, making it well-suited for rapid diagnosis of known pathogens and resistance genes. However, its reliance on a predefined panel limits its scope. In contrast, NMgS enabled broad, untargeted detection of pathogens and AMR genes, but its sensitivity was hindered by residual host DNA and the lack of a universally optimized depletion protocol. While longer sequencing times or reduced sample throughput could improve NMgS performance, these adjustments are impractical for routine clinical use. Overall, NTS offers advantages in speed, cost, and sensitivity for acute infections, whereas NMgS is better suited for complex or atypical cases involving unknown or novel pathogens. Importantly, sequencing-based diagnostics should complement rather than replace traditional culture, which remains essential for confirming phenotypic resistance and detecting pathogens in low-biomass samples.

## Supporting information

Supplemental 3: The concordance between NTS and culture in pathogen identification

Supplemental Table 1. Primer sequences of ID and AMR primer set

Supplemental 4: The concordance between NMgS and culture in pathogen identification

Supplemental 1. Detailed protocol for NTS workflow

Supplemental 2. Detailed protocols for NMgS workflow

## Data Availability

All sequencing data generated in this study are publicly available through the NCBI BioProject database under accession numbers PRJNA1270715 and PRJNA1270737.

