## Supplemental 3: The concordance between NTS and culture in pathogen identification for "Optimizing and Evaluating Nanopore-Based Targeted and Metagenomic Sequencing Workflows for Rapid Diagnosis of Acute Invasive Infections from Normally Sterile Body Fluids"

| **Supplemental 3: The concordance between NTS and culture in pathogen identification** | | | | | |  |  |  |
| --- | --- | --- | --- | --- | --- | --- | --- | --- |
| **No.** | **Sample ID** | **No. of cultured species** | **Culture ID** | **16S/ ITS ID** | **Read count** | **Relative abundance** | **Total classified reads** | **Concordance** |
| 1 | 23MB047690 | monomicrobial | Staphylococcus aureus (MRSA) (scanty) | Staphylococcus aureus | 825 | 0.952 | 867 | matched |
| 2 | 23MB047738 | monomicrobial | Streptococcus agalactiae (heavy) | Streptococcus agalactiae/ Streptococcus sp. FDAARGOS_520/ Streptococcus sp. FDAARGOS_522 | 8381 | 1 | 8381 | matched |
| 3 | 23MB047824 | monomicrobial | Streptococcus pyogenes (moderate) | Streptococcus pyogenes | 10409 | 1 | 10409 | matched |
| 4 | 23MB047892 | monomicrobial | Staphylococcus aureus (heavy) | Staphylococcus aureus | 97 | 1 | 97 | matched |
| 5 | 23MB087252 | monomicrobial | Escherichia coli (scanty) | Not found | 0 | 0 | 28 | unmatched |
| 6 | 23MB087291 | monomicrobial | Candida albicans (Scanty) | Candida africana (C. albicans complex) | 9 | 0.243 | 37 | matched |
| 7 | 23MB087709 | monomicrobial | Klebsiella pneumoniae complex (heavy) | Klebsiella pneumoniae | 12392 | 1 | 12392 | matched |
| 8 | 23MB087820 | monomicrobial | Klebsiella pneumoniae complex (scanty) | Klebsiella pneumoniae | 243 | 0.469 | 518 | matched |
| 9 | 23MB087882 | monomicrobial | Staphylococcus aureus (scanty) | Staphylococcus aureus | 1330 | 1 | 1330 | matched |
| 10 | 23MB101355 | monomicrobial | Bacteroides fragilis (moderate) | Bacteroides fragilis | 219 | 1 | 219 | matched |
| 11 | 23MB101356 | monomicrobial | Staphylococcus aureus (heavy) | Staphylococcus aureus | 1788 | 1 | 1788 | matched |
| 12 | 23MB101447 | monomicrobial | Streptococcus dysgalactiae subsp. equisimilis (heavy) | Streptococcus dysgalactiae | 5665 | 1 | 5665 | matched |
| 13 | 23MB101616 | monomicrobial | Staphylococcus aureus (scanty) | Staphylococcus aureus | 12853 | 0.999 | 12863 | matched |
| 14 | 23MB110604 | monomicrobial | Campylobacter fetus (isolated) | Campylobacter fetus | 2 | 0.08 | 25 | matched |
| 15 | 23MB110726 | monomicrobial | Corynebacterium striatum (scanty) | Corynebacterium striatum | 5 | 0.625 | 8 | matched |
| 16 | 23M2010586 | polymicrobial | Enterococcus faecium (Heavy) | Enterococcus faecium | 27088 | 0.874 | 30977 | matched |
|  |  |  | Escherichia coli (Scanty) | Escherichia coli | 54 | 0.002 |  | matched |
|  |  |  | Klebsiella variicola (Scanty) | Klebsiella pneumoniae | 46 | 0.001 |  | matched |
|  |  |  | Enterococcus raffinosus (Moderate) | Enterococcus gilvus | 880 | 0.028 |  | matched |
|  |  |  | Clostridium perfringens (Scanty) | Clostridium perfringens | 232 | 0.007 |  | matched |
|  |  |  | Alpha-haemolytic Streptococci (Scanty) | Streptococcus oralis | 115 | 0.004 |  | matched |
|  |  |  | Proteus species (Scanty) | Proteus vulgaris | 17 | 0.001 |  | matched |
|  |  |  | Streptococcus infantarius (Scanty) | Streptococcus infantarius | 438 | 0.014 |  | matched |
| 17 | 23M2010695 | polymicrobial | Escherichia coli (Heavy) | Escherichia coli | 365 | 0.009 | 42083 | matched |
|  |  |  | Enterococcus faecalis (Moderate) | Enterococcus faecalis | 36050 | 0.857 |  | matched |
|  |  |  | Enterococcus avium (Scanty) | Enterococcus avium | 5417 | 0.129 |  | matched |
| 18 | 23M2010728 | polymicrobial | Escherichia coli (Moderate) | Escherichia coli | 18819 | 0.837 | 22472 | matched |
|  |  |  | Proteus mirabilis (Moderate) | Proteus mirabilis | 19 | 0.001 |  | matched |
|  |  |  | Klebsiella pneumoniae (Scanty) | Klebsiella pneumoniae | 30 | 0.001 |  | matched |
|  |  |  | Clostridium perfringens (Scanty) | Clostridium perfringens | 3491 | 0.155 |  | matched |
| 19 | 23M2011494 | polymicrobial | Escherichia coli (Scanty) | Escherichia coli | 15459 | 0.279 | 55430 | matched |
|  |  |  | Bacteroides fragilis (Scanty) | Bacteroides fragilis | 3151 | 0.057 |  | matched |
|  |  |  | Coagulase negative Staphylococcus (Scanty) | Not found | 0 | 0 |  | unmatched |
|  |  |  | Streptococcus constellatus (Scanty) | Not found | 0 | 0 |  | unmatched |
| 20 | 23M2011587 | polymicrobial | Corynebacterium aurimucosum (Moderate) | Corynebacterium aurimucosum | 379 | 0.007 | 53824 | matched |
|  |  |  | Coagulase negative Staphylococcus (Heavy) | Staphylococcus epidermidis | 34826 | 0.647 |  | matched |
| 21 | 23M2102729 | polymicrobial | Escherichia coli (Heavy) | Escherichia coli | 5546 | 0.276 | 20114 | matched |
|  |  |  | Pseudomonas aeruginosa (Scanty) | Pseudomonas aeruginosa | 49 | 0.002 |  | matched |
|  |  |  | Streptococcus constellatus (Scanty) | Streptococcus constellatus | 23 | 0.001 |  | matched |
| 22 | 23M2102735 | monomicrobial | Candida albicans (Scanty) | Candida africana (C. albicans complex) | 173 | 0.416 | 416 | matched |
| 23 | 23M2102736 | monomicrobial | Escherichia coli | Escherichia coli | 18585 | 0.982 | 18916 | matched |
| 24 | 23M2102738 | polymicrobial | Escherichia coli (Moderate) | Escherichia coli | 718 | 0.027 | 26997 | matched |
|  |  |  | Acinetobacter calcoaceticus-baumannii complex (Heavy) | Acinetobacter baumannii | 91 | 0.003 |  | matched |
|  |  |  | Klebsiella pneumoniae (Scanty) | Klebsiella pneumoniae | 13 | 0.0005 |  | matched |
|  |  |  | Lysinibacillus species (Scanty) | Lysinibacillus sp. YS11 | 102 | 0.004 |  | matched |
| 25 | 23M2102747 | monomicrobial | Coagulase negative Staphylococcus | Staphylococcus capitis | 64 | 0.075 | 860 | matched |
| 26 | 23M2102754 | monomicrobial | Bacteroides pyogenes (Heavy) | Bacteroides pyogenes | 33039 | 0.837 | 39463 | matched |
| 27 | 23M2102780 | monomicrobial | Enterococcus gallinarum | Not found | 0 | 0 | 177 | unmatched |
| 28 | 23M2102832 | monomicrobial | Staphylococcus aureus (MRSA) (Heavy) | Staphylococcus aureus | 28855 | 1 | 28855 | matched |
| 29 | 23M2102908 | monomicrobial | Bacteroides fragilis | Bacteroides fragilis | 9 | 0.01 | 872 | matched |
| 30 | 23M4201725 | monomicrobial | Streptococcus intermedius (Heavy) | Streptococcus intermedius | 67 | 1 | 67 | matched |
| 31 | 23M2102914 | monomicrobial | Staphylococcus aureus (MRSA) | Staphylococcus aureus | 3554 | 0.972 | 3658 | matched |
| 32 | 23M2102931 | monomicrobial | Pseudomonas aeruginosa (Heavy) | Pseudomonas aeruginosa | 7155 | 1 | 7155 | matched |
| 33 | 23M9006507 | monomicrobial | Staphylococcus aureus (Moderate) | Staphylococcus aureus | 215 | 0.935 | 230 | matched |
| 34 | 23M2104323 | polymicrobial | Escherichia coli (Heavy) | Not found | 0 | 0 | 2261 | unmatched |
|  |  |  | Bacteroides fragilis (Heavy) | Bacteroides fragilis | 229 | 0.101 |  | matched |
|  |  |  | Enterococcus faecalis (Scanty) | Not found | 0 | 0 |  | unmatched |
| 35 | 23M2104297 | monomicrobial | Staphylococcus aureus | Staphylococcus aureus | 47 | 0.58 | 81 | matched |
| 36 | 23M2104285 | monomicrobial | Acinetobacter calcoaceticus-baumannii complex (Moderate) | Acinetobacter pittii | 1973 | 1 | 1973 | matched |
| 37 | 23M2104292 | monomicrobial | Coagulase negative Staphylococcus | Staphylococcus epidermidis | 7 | 0.233 | 30 | matched |
| 38 | 23M2104271 | monomicrobial | Streptococcus mitis group (Scanty) | Not found | 0 | 0 | 16 | unmatched |
| 39 | 23M2104267 | monomicrobial | Escherichia coli (Scanty) | Escherichia coli | 1516 | 1 | 1516 | matched |
| 40 | 23M2104266 | polymicrobial | Candida albicans (Scanty) | Candida africana (C. albicans complex) | 29 | 0.42 | 69 | matched |
|  |  |  | Candida tropicalis (Scanty) | Not found | 0 | 0 |  | unmatched |
| 41 | 23M2104242 | monomicrobial | Escherichia coli (Scanty) | Escherichia coli | 60 | 0.035 | 1728 | matched |
| 42 | 23M2104135 | monomicrobial | Candida parapsilosis | Not found | 0 | 0 | 6 | unmatched |
| 43 | 23M2104041 | monomicrobial | Coagulase negative Staphylococcus | Staphylococcus hominis | 3 | 0.111 | 27 | matched |
| 44 | 23M2104033 | monomicrobial | Streptococcus anginosus | Streptococcus anginosus | 6 | 0.207 | 29 | matched |
| 45 | 23M2103978 | monomicrobial | Streptococcus dysgalactiae (Scanty) | Streptococcus dysgalactiae | 5305 | 1 | 5305 | matched |
| 46 | 23M2103952 | monomicrobial | Corynebacterium striatum | Not found | 0 | 0 | 39 | unmatched |
| 47 | 23M2103944 | monomicrobial | Escherichia coli (Heavy) | Escherichia coli | 4372 | 1 | 4372 | matched |
| 48 | 23M2104372 | monomicrobial | Staphylococcus aureus (Scanty) | Staphylococcus aureus | 126 | 1 | 126 | matched |
| 49 | 23M2104408 | monomicrobial | Streptococcus parasanguinis | Streptococcus parasanguinis | 192 | 0.98 | 196 | matched |
| 50 | 23M2104413 | polymicrobial | Escherichia coli (Moderate) | Not found | 0 | 0 | 857 | unmatched |
|  |  |  | Peptostreptococcus anaerobius (Scanty) | Peptostreptococcus anaerobius | 36 | 0.042 |  | matched |
|  |  |  | Actinomyces turicensis (Scanty) | Not found | 0 | 0 |  | unmatched |
|  |  |  | Prevotella intermedia (Moderate) | Prevotella intermedia | 6 | 0.007 |  | matched |
|  |  |  | Bacteroides cellulosilyticus (Moderate) | Not found | 0 | 0 |  | unmatched |
|  |  |  | Bacteroides ovatus (Scanty) | Not found | 0 | 0 |  | unmatched |
| 51 | 23M2104421 | monomicrobial | Staphylococcus lugdunensis (Scanty) | Not found | 0 | 0 | 2 | unmatched |
| 52 | 23M2104412 | polymicrobial | Streptococcus anginosus (Scanty) | Not found | 0 | 0 | 2921 | unmatched |
|  |  |  | Rothia mucilaginosa (Scanty) | Rothia mucilaginosa | 125 | 0.043 |  | matched |
|  |  |  | Streptococcus mitis group (Scanty) | Streptococcus mitis | 227 | 0.078 |  | matched |
|  |  |  | Staphylococcus aureus | Not found | 0 | 0 |  | unmatched |
|  |  |  | Streptococcus vestibularis | Streptococcus sp. FDAARGOS_192 | 126 | 0.043 |  | matched |
|  |  |  | Lactobacillus salivarius | Not found | 0 | 0 |  | unmatched |
| 53 | 24MB001633 | monomicrobial | Staphylococcus species (isolated) | Staphylococcus aureus | 33 | 1 | 33 | matched |
| 54 | 24MB011005 | polymicrobial | Streptococcus agalactiae (Group B streptococcus) (scanty) | Not found | 0 | 0 | 16 | unmatched |
|  |  |  | Viridans streptococcus (heavy) | Streptococcus oralis | 3 | 0.188 |  | matched |
| 55 | 24MB011006 | monomicrobial | Staphylococcus aureus (MRSA) (scanty) | Staphylococcus aureus | 5 | 0.833 | 6 | matched |
| 56 | 24MB011017 | monomicrobial | Klebsiella pneumoniae complex (scanty) | Klebsiella pneumoniae | 98 | 0.97 | 101 | matched |
| 57 | 24MB011020 | monomicrobial | Enterococcus faecalis (moderate) | Enterococcus faecalis | 504 | 1 | 504 | matched |
| 58 | 24MB011073 | polymicrobial | Staphylococcus aureus (scanty) | Staphylococcus aureus | 280 | 1 | 280 | matched |
|  |  |  | Streptococcus agalactiae (scanty) | Not found | 0 | 0 |  | unmatched |
| 59 | 24MB011062 | polymicrobial | Escherichia coli (scanty) | Escherichia coli | 29 | 0.242 | 120 | matched |
|  |  |  | Enterococcus faecium (isolated) | Not found | 0 | 0 |  | unmatched |
| 60 | 24MB011066 | monomicrobial | Pseudomonas aeruginosa (heavy) | Pseudomonas aeruginosa | 80 | 1 | 80 | matched |
| 61 | 24MB011104 | polymicrobial | Streptococcus anginosus group (heavy) | Streptococcus intermedius | 124 | 0.139 | 889 | matched |
|  |  |  | Parvimonas micra (heavy) | Parvimonas micra | 760 | 0.855 |  | matched |
| 62 | 24MB011155 | polymicrobial | Escherichia coli (heavy) | Not found | 0 | 0 | 207 | unmatched |
|  |  |  | Enterococcus faecium (heavy) | Enterococcus faecium | 207 | 1 |  | matched |
|  |  |  | Acinetobacter baumannii complex (scanty) | Not found | 0 | 0 |  | unmatched |
|  |  |  | Burkholderia cepacia complex (scanty) | Not found | 0 | 0 |  | unmatched |
|  |  |  | Candida albicans (scanty) | Candida africana | 3 | 0.103 | 29 | matched |
| 63 | 24MB011121 | polymicrobial | Klebsiella species (moderate) | Klebsiella michiganensis | 4 | 0.007 | 604 | matched |
|  |  |  | Escherichia coli (heavy) | Escherichia coli | 16 | 0.026 |  | matched |
|  |  |  | Enterococcus raffinosus (heavy) | Enterococcus gilvus | 383 | 0.634 |  | matched |
|  |  |  | Clostridium perfringes (scanty) | Clostridium perfringens | 46 | 0.076 |  | matched |
|  |  |  | Pseudomonas aeruginosa (moderate) | Not found | 0 | 0 |  | unmatched |
| 64 | 24MB011122 | monomicrobial | Escherichia coli (heavy) | Escherichia coli | 248 | 1 | 248 | matched |
| 65 | 24MB011188 | monomicrobial | Escherichia coli (heavy) | Escherichia coli | 13 | 1 | 13 | matched |
| 66 | 24MB011103 | monomicrobial | Escherichia coli (scanty) | Escherichia coli | 300 | 0.993 | 302 | matched |
| 67 | 24MB011255 | monomicrobial | Pseudomonas aeruginosa (scanty) | Pseudomonas aeruginosa | 34 | 1 | 34 | matched |
| 68 | 24MB011363 | polymicrobial | Escherichia coli (heavy) | Escherichia coli | 126 | 0.813 | 155 | matched |
|  |  |  | Proteus mirabilis (heavy) | Proteus mirabilis | 25 | 0.161 |  | matched |
|  |  |  | Candida tropicalis (scanty) | Not found | 0 | 0 | 0 | unmatched |
| 69 | 24MB011403 | polymicrobial | Escherichia coli (heavy) | Escherichia coli | 24 | 0.162 | 148 | matched |
|  |  |  | Enterococcus faecalis (heavy) | Enterococcus faecalis | 124 | 0.838 |  | matched |
|  |  |  | Klebsiella oxytoca (heavy) | Not found | 0 | 0 |  | unmatched |
| 70 | 24MB011364 | polymicrobial | Klebsiella pneumoniae complex (heavy) | Klebsiella pneumoniae | 1347 | 1 | 1347 | matched |
|  |  |  | Pseudomonas aeruginosa (heavy) | Not found | 0 | 0 |  | unmatched |
| 71 | 24MB011412 | monomicrobial | Escherichia coli (heavy) | Escherichia coli | 82 | 0.965 | 85 | matched |
| 72 | 24MB011541 | monomicrobial | Viridans streptococcus (heavy) | Streptococcus gordonii | 1798 | 1 | 1798 | matched |
| 73 | 24MB011567 | monomicrobial | Phocaeicola vulgatus (heavy) | Phocaeicola vulgatus | 545 | 1 | 545 | matched |
| 74 | 24MB011665 | monomicrobial | Klebsiella pneumoniae complex (moderate) | Not found | 0 | 0 | 0 | unmatched |
| 75 | 24MB011666 | monomicrobial | Staphylococcus aureus (heavy) | Staphylococcus aureus | 141 | 1 | 141 | matched |
| 76 | 24MB011667 | monomicrobial | Klebsiella pneumoniae complex (heavy) | Klebsiella pneumoniae | 101 | 0.99 | 102 | matched |
| 77 | 24MB011662 | monomicrobial | Pseudomonas aeruginosa (scanty) | Pseudomonas aeruginosa | 23 | 1 | 23 | matched |
| 78 | 24MB011753 | monomicrobial | Klebsiella aerogenes (heavy) | Klebsiella aerogenes | 187 | 1 | 187 | matched |
| 79 | 24MB011679 | polymicrobial | Acinetobacter baumannii complex (scanty) | Not found | 0 | 0 | 1798 | unmatched |
|  |  |  | Staphylococcus aureus (MRSA) (moderate) | Staphylococcus aureus | 129 | 0.072 |  | matched |
|  |  |  | Enterococcus casseliflavus (heavy) | Enterococcus sp. FDAARGOS_375 | 1066 | 0.593 |  | matched |
|  |  |  | Lactobacillus paracasei (heavy) | Lactobacillus paracasei | 590 | 0.328 |  | matched |
|  |  |  | Candida tropicalis (heavy) | Candida tropicalis | 2028 | 1 | 2029 | matched |
| 80 | 24MB011678 | polymicrobial | Enterobacter species (heavy) | Enterobacter roggenkampii | 785 | 0.044 | 17780 | matched |
|  |  |  | Kluyvera georgiana (moderate) | Not found | 0 | 0 |  | unmatched |
|  |  |  | GPC in chains (heavy) | Streptococcus pasteurianus | 15262 | 0.858 |  | matched |
| 81 | 24MB011637 | polymicrobial | Pseudomonas aeruginosa (heavy) | Pseudomonas aeruginosa | 261 | 0.154 | 1696 | matched |
|  |  |  | Escherichia coli (moderate) | Escherichia coli | 723 | 0.426 |  | matched |
|  |  |  | Klebsiella pneumoniae complex (heavy) | Not found | 0 | 0 |  | unmatched |
|  |  |  | Bacteroides species (heavy) | Not found | 0 | 0 |  | unmatched |
|  |  |  | Enterococcus faecium (scanty) | Not found | 0 | 0 |  | unmatched |
| 82 | 23MB170626 | monomicrobial | Pseudomonas aeruginosa (isolated) | Pseudomonas aeruginosa | 123 | 0.75 | 164 | matched |
| 83 | 23MB170293 | monomicrobial | Staphylococcus aureus (scanty) | Staphylococcus aureus | 6556 | 1 | 6556 | matched |
| 84 | 23MB184156 | monomicrobial | Staphylococcus aureus (moderate) | Staphylococcus aureus | 11294 | 1 | 11294 | matched |
| 85 | 23MB184024 | monomicrobial | Staphylococcus aureus (MRSA) (moderate) | Staphylococcus aureus | 628 | 0.998 | 629 | matched |
| 86 | 23MB170776 | polymicrobial | Pseudomonas aeruginosa (heavy) | Pseudomonas aeruginosa | 20270 | 1 | 20270 | matched |
|  |  |  | Klebsiella pneumoniae complex (isolated) | Not found | 0 | 0 |  | unmatched |
| 87 | 23MB170782 | monomicrobial | Staphylococcus aureus (MRSA) (scanty) | Staphylococcus aureus | 64 | 0.97 | 66 | matched |
| 88 | 23MB170774 | monomicrobial | Streptococcus anginosus group (scanty) | Streptococcus intermedius | 10345 | 0.707 | 14638 | matched |
| 89 | 23MB170490 | polymicrobial | Bacteroides fragilis (moderate) | Bacteroides fragilis | 1912 | 0.834 | 2292 | matched |
|  |  |  | Escherichia coli (moderate) | Escherichia coli | 380 | 0.166 |  | matched |
| 90 | 23MB170331 | monomicrobial | Aeromonas species (heavy) | Aeromonas veronii | 8869 | 0.855 | 10379 | matched |
|  |  |  |  | Aeromonas jandaei | 1510 | 0.145 |  |  |
| 91 | 23MB184255 | polymicrobial | Streptococcus anginosus group (heavy) | Streptococcus anginosus | 92 | 0.015 | 6314 | matched |
|  |  |  | Escherichia coli (heavy) | Not found | 0 | 0 |  | unmatched |
|  |  |  | Bacteroides thetaiotaomicron (heavy) | Bacteroides thetaiotaomicron | 121 | 0.019 |  | matched |
| 92 | 23MB184185 | monomicrobial | Streptococcus anginosus group (scanty) | Streptococcus intermedius | 823 | 0.959 | 858 | matched |
| 93 | 24MB017342 | polymicrobial | Klebsiella pneumoniae complex (moderate) | Klebsiella pneumoniae | 8 | 0.018 | 454 | matched |
|  |  |  | Escherichia coli (heavy) | Escherichia coli | 185 | 0.407 |  | matched |
|  |  |  | Enterococcus avium (heavy) | Enterococcus avium | 246 | 0.542 |  | matched |
| 94 | 24MB017347 | polymicrobial | Escherichia coli (moderate) | Escherichia coli | 1284 | 0.07 | 18443 | matched |
|  |  |  | Aeromonas species (heavy) | Aeromonas caviae | 8805 | 0.477 |  | matched |
|  |  |  | Klebsiella oxytoca (heavy) | Klebsiella oxytoca | 224 | 0.012 |  | matched |
|  |  |  | Enterococcus species (scanty) | Enterococcus gilvus | 374 | 0.02 |  | matched |
|  |  |  | Proteus species (scanty) | Proteus vulgaris | 101 | 0.005 |  | matched |
| 95 | 24MB017428 | monomicrobial | Klebsiella pneumoniae complex (scanty) | Klebsiella pneumoniae | 15 | 0.081 | 186 | matched |
| 96 | 24MB017427 | monomicrobial | Escherichia coli (heavy) | Escherichia coli | 677 | 0.999 | 678 | matched |
| 97 | 24MB017494 | monomicrobial | Escherichia coli (moderate) | Not found | 0 | 0 | 2896 | unmatched |
| 98 | 24MB017555 | polymicrobial | Escherichia coli (heavy) | Escherichia coli | 191 | 0.01 | 18233 | matched |
|  |  |  | Streptococcus anginosus group (moderate) | Streptococcus anginosus | 75 | 0.004 |  | matched |
|  |  |  | Bacteroides species (heavy) | Bacteroides fragilis | 547 | 0.03 |  | matched |
|  |  |  |  | Bacteroides thetaiotaomicron | 245 | 0.013 |  |  |
| 99 | 24MB017777 | polymicrobial | Streptococcus anginosus group (scanty) | Not found | 0 | 0 | 547 | unmatched |
|  |  |  | Peptoniphilus species (scanty) | Peptoniphilus harei | 98 | 0.179 |  | matched |
|  |  |  |  | Peptoniphilus duerdenii | 23 | 0.042 |  |  |
| 100 | 24M2101353 | polymicrobial | Streptococcus vestibularis | Streptococcus sp. FDAARGOS_192 / Streptococcus vestibularis | 84 | 0.977 | 86 | matched |
|  |  |  | Streptococcus salivarius | Not found | 0 | 0 |  | unmatched |
| 101 | 24M2101323 | monomicrobial | Coagulase negative Staphylococcus (scanty) | Staphylococcus pasteuri | 1618 | 1 | 1618 | matched |
| 102 | 24M2004435 | monomicrobial | Escherichia coli (moderate) | Escherichia coli | 17603 | 1 | 17603 | matched |
| 103 | 24M2004419 | polymicrobial | Escherichia coli (moderate) | Escherichia coli | 3542 | 0.822 | 4309 | matched |
|  |  |  | Enterococcus hirae (moderate) | Enterococcus hirae | 15 | 0.003 |  | matched |
|  |  |  | Clostridium perfringens (moderate) | Not found | 0 | 0 |  | unmatched |
| 104 | 24MB032054 | monomicrobial | Pseudomonas species | Pseudomonas monteilii | 20897 | 0.992 | 21056 | matched |
| 105 | 24MB032068 | polymicrobial | Acinetobacter baumannii complex | Acinetobacter baumannii | 5524 | 0.99 | 5582 | matched |
|  |  |  | candida tropicalis | candida tropicalis | 8134 | 0.997 | 8157 | matched |
| 106 | 24MB032078 | monomicrobial | Viridans Streptococcus | Streptococcus salivarius | 5658 | 0.994 | 5692 | matched |
| 107 | 24MB032085 | monomicrobial | Pseudomonas species (isolated) | Pseudomonas monteilii | 91 | 0.181 | 500 | matched |
| 108 | 24MB032086 | polymicrobial | Escherichia coli (heavy) | Escherichia coli | 292 | 0.012 | 24984 | matched |
|  |  |  | Pseudomonas aeruginosa (moderate) | Pseudomonas aeruginosa | 14 | 0.001 |  | matched |
|  |  |  | Streptococcus anginosus group (heavy) | Streptococcus intermedius | 3506 | 0.14 |  | matched |
|  |  |  | Bacteroides thetaiotaomicron (heavy) | Bacteroides thetaiotaomicron | 48 | 0.002 |  | matched |
| 109 | 24MB110434 | monomicrobial | Klebsiella pneumoniae complex (scanty) | Klebsiella pneumoniae | 2059 | 1 | 2059 | matched |
| 110 | 24MB110571 | monomicrobial | Staphylococcus aureus (heavy) | Staphylococcus aureus | 2513 | 1 | 2513 | matched |
| 111 | 24MB110656 | monomicrobial | Staphylococcus aureus (moderate) | Staphylococcus aureus | 3546 | 1 | 3546 | matched |
| 112 | 24MB110723 | monomicrobial | Citrobacter koseri (scanty) | Citrobacter koseri | 8062 | 0.999 | 8074 | matched |
| 113 | 24MB111063 | monomicrobial | Enterococcus faecalis (moderate) | Enterococcus faecalis | 8 | 0.421 | 19 | matched |
| 114 | 24MB111194 | polymicrobial | Pseudomonas aeruginosa (moderate) | Pseudomonas aeruginosa | 6023 | 0.376 | 16022 | matched |
|  |  |  | Morganella morganii (moderate) | Morganella morganii | 9597 | 0.599 |  | matched |
| 115 | 24MB111565 | polymicrobial | Morganella morganii (moderate) | Morganella morganii | 5748 | 0.924 | 6219 | matched |
|  |  |  | Escherichia coli (scanty) | Escherichia coli | 471 | 0.076 |  | matched |
| 116 | 24MB111308 | monomicrobial | Staphylococcus aureus (moderate) | Staphylococcus aureus | 3987 | 1 | 3987 | matched |
| 117 | 24MB111311 | monomicrobial | Citrobacter freundii complex (moderate) | Citrobacter freundii complex | 229 | 0.094 | 2442 | matched |
| 118 | 24MB111315 | polymicrobial | Escherichia coli (moderate) | Escherichia coli | 14 | 0.001 | 13365 | matched |
|  |  |  | Streptococcus constellatus (scanty) | Not found | 0 | 0 |  | unmatched |
|  |  |  | Parvimonas micra (heavy) | Parvimonas micra | 7986 | 0.598 |  | matched |
| 119 | 24MB111480 | monomicrobial | Staphylococcus aureus (moderate) | Staphylococcus aureus | 48599 | 1 | 48599 | matched |
| 120 | 24MB111876 | monomicrobial | Klebsiella pneumoniae complex (heavy) | Klebsiella pneumoniae | 9444 | 1 | 9444 | matched |
| 121 | 24MB111852 | monomicrobial | Klebsiella pneumoniae complex (moderate) | Not found | 0 | 0 | 0 | unmatched |
| 122 | 24MB112057 | monomicrobial | Citrobacter koseri (scanty) | Citrobacter koseri | 7397 | 1 | 7397 | matched |
| 123 | 24MB112341 | polymicrobial | Acinetobacter calcoaceticus baumannii complex (scanty) | Acinetobacter baumannii | 119 | 0.056 | 2114 | matched |
|  |  |  | Staphylococcus aureus (MRSA) (scanty) | Staphylococcus aureus | 1995 | 0.944 |  | matched |
|  |  |  | Bacteroides species (scanty) | Not found | 0 | 0 |  | unmatched |
| 124 | 24MB112432 | monomicrobial | Pseudomonas aeruginosa (scanty) | Pseudomonas aeruginosa | 503 | 0.997 | 505 | matched |
| 125 | 24MB112433 | monomicrobial | Pseudomonas aeruginosa (scanty) | Pseudomonas aeruginosa | 2173 | 0.967 | 2248 | matched |
| 126 | 24MB112478 | monomicrobial | Escherichia coli (scanty) | Escherichia coli | 4.67 | 0.667 | 7 | matched |
| 127 | 24MB111695 | monomicrobial | Campylobacter species (scanty) | Campylobacter fetus | 265 | 1 | 265 | matched |
| 128 | 24MB111720 | monomicrobial | Streptococcus anginosus (moderate) | Streptococcus milleri | 9527 | 0.736 | 12942 | matched |
| 129 | 24MB113274 | monomicrobial | Klebsiella pneumoniae complex (moderate) | Klebsiella pneumoniae | 3737 | 1 | 3737 | matched |
| 130 | 24MB112605 | monomicrobial | Klebsiella pneumoniae complex (scanty) | Klebsiella pneumoniae | 2845 | 0.326 | 8733 | matched |
| 131 | 24MB112927 | monomicrobial | Streptococcus milleri group (scanty) | Streptococcus intermedius | 8608 | 0.585 | 14705 | matched |
| 132 | 24MB113276 | polymicrobial | Morganella morganii (heavy) | Morganella morganii | 1453 | 0.051 | 28586 | matched |
|  |  |  | Streptococcus constellatus (scanty) | Not found | 0 | 0 |  | unmatched |
|  |  |  | Fusobacterium mortiferum (scanty) | Fusobacterium mortiferum | 187 | 0.007 |  | matched |
|  |  |  | Clostridium ramosum (Thomasclavelia ramosa) (scanty) | Not found | 0 | 0 |  | unmatched |
| 133 | 24MB113346 | monomicrobial | Escherichia coli (scanty) | Escherichia coli | 586 | 0.094 | 6207 | matched |
| 134 | 24MB113395 | monomicrobial | Klebsiella pneumoniae complex (moderate) | Klebsiella variicola | 5875 | 0.961 | 6114 | matched |
| 135 | 24MB113404 | monomicrobial | Streptococcus intermedius (moderate) | Streptococcus intermedius | 6083 | 1 | 6083 | matched |
| 136 | 24MB113415 | monomicrobial | Streptococcus agalactiae (scanty) | Streptococcus agalactiae | 1362 | 1 | 1362 | matched |
| 137 | 24MB113419 | polymicrobial | Escherichia coli (scanty) | Escherichia coli | 738 | 0.114 | 6484 | matched |
|  |  |  | Klebsiella pneumoniae complex (scanty) | Klebsiella pneumoniae | 790 | 0.122 |  | matched |
| 138 | 24MB113466 | monomicrobial | Klebsiella pneumoniae complex (scanty) | Klebsiella pneumoniae | 4402 | 1 | 4402 | matched |
| 139 | 24MB117350 | monomicrobial | Streptococcus constellatus (heavy) | Streptococcus intermedius | 311 | 0.26 | 1196 | matched |
| 140 | 24MB117532 | monomicrobial | Salmonella species, Group D (moderate) | Salmonella enterica | 1760 | 1 | 1760 | matched |
| 141 | 24MB117763 | monomicrobial | Klebsiella pneumoniae complex (scanty) | Klebsiella pneumoniae | 9 | 0.643 | 14 | matched |
| 142 | 24MB117992 | polymicrobial | Escherichia coli (moderate) | Escherichia coli | 33 | 0.005 | 7054 | matched |
|  |  |  | Enterococcus raffinosus (heavy) | Not found | 0 | 0 |  | unmatched |
| 143 | 24MB118151 | polymicrobial | Escherichia coli (moderate) | Escherichia coli | 507 | 0.091 | 5554 | matched |
|  |  |  | Klebsiella oxytoca (moderate) | Not found | 0 | 0 |  | unmatched |
|  |  |  | Enterococcus avium (scanty) | Not found | 0 | 0 |  | unmatched |
| 144 | 24MB118533 | polymicrobial | Streptococcus anginosus (heavy) | Streptococcus milleri | 3684 | 0.467 | 7895 | matched |
|  |  |  | Raoutella ornithinolytica (heavy) | Raoultella ornithinolytica | 3324 | 0.421 |  | matched |
|  |  |  | Bacteroides fragilis (scanty) | Bacteroides fragilis | 343 | 0.044 |  | matched |
| 145 | 24MB119459 | monomicrobial | Klebsiella pneumoniae complex (heavy) | Klebsiella pneumoniae | 37 | 1 | 37 | matched |
| 146 | 24MB119528 | polymicrobial | Escherichia coli (heavy) | Escherichia coli | 655 | 0.067 | 9799 | matched |
|  |  |  | Streptococcus anginosus (scanty) | Not found | 0 | 0 |  | unmatched |
|  |  |  | Bacteriodes caccae (heavy) | Bacteroides caccae | 66 | 0.007 |  | matched |
|  |  |  | Clostridium perfringens (scanty) | Clostridium perfringens | 130 | 0.013 |  | matched |
| 147 | 24MB120055 | monomicrobial | Escherichia coli (scanty) | Escherichia coli | 2 | 0.003 | 537 | matched |
| 148 | 24MB120463 | monomicrobial | Streptococcus agalactiae (scanty) | Streptococcus agalactiae | 1205 | 1 | 1205 | matched |
| 149 | 24MB120515 | polymicrobial | Escherichia coli (moderate) | Escherichia coli | 23 | 0.004 | 6360 | matched |
|  |  |  | Enterococcus avium (scanty) | Enterococcus avium | 13 | 0.002 |  | matched |
|  |  |  | Clostridium innocuum (scanty) | Not found | 0 | 0 |  | unmatched |
|  |  |  | Peptostreptococcus anaerobius (scanty) | Peptostreptococcus anaerobius | 1286 | 0.202 |  | matched |
|  |  |  | Bacteriodes fragilis (heavy) | Bacteroides fragilis | 4583 | 0.721 |  | matched |
| 150 | 24MB120576 | monomicrobial | Staphylococcus aureus (moderate) | Staphylococcus aureus | 1547 | 1 | 1547 | matched |
| 151 | 24MB122216 | monomicrobial | Staphylococcus aureus (MRSA) (heavy) | Staphylococcus aureus | 20012 | 1 | 20012 | matched |
| 152 | 24MB122452 | polymicrobial | Neisseria flavescens subflava group (moderate) | Neisseria subflava | 8 | 0.055 | 146 | matched |
|  |  |  | Streptococcus mitis group (moderate) | Streptococcus anginosus | 29 | 0.196 |  | matched |
|  |  |  | Candia glabrata (heavy) | Nakaseomyces glabratus | 586 | 0.998 | 587 | matched |
| 153 | 24MB122527 | polymicrobial | Escherichia coli (moderate) | Not found | 0 | 0 | 0 | unmatched |
|  |  |  | Citrobacter koseri (moderate) | Not found | 0 | 0 |  | unmatched |
|  |  |  | Enterococcus raffinosus (moderate) | Not found | 0 | 0 |  | unmatched |
|  |  |  | Candida glabrata (heavy) | Nakaseomyces glabratus | 102 | 0.962 | 106 | matched |
| 154 | 24MB122856 | monomicrobial | Salmonella species, Group B (moderate) | Salmonella enterica | 508 | 0.998 | 509 | matched |
| 155 | 24MB122862 | monomicrobial | Klebsiella pneumoniae complex (scanty) | Klebsiella pneumoniae | 112 | 0.957 | 117 | matched |
| 156 | 24MB123950 | monomicrobial | Streptococcus intermedius (moderate) | Streptococcus intermedius | 6625 | 1 | 6625 | matched |
| 157 | 24MB123988 | monomicrobial | Pseudomonas aeruginosa (scanty) | Pseudomonas aeruginosa | 4140 | 1 | 4140 | matched |
| 158 | 24MB124304 | monomicrobial | Streptococcus intermedius (moderate) | Streptococcus intermedius | 4276 | 1 | 4276 | matched |
| 159 | 24MB125173 | polymicrobial | Escherichia coli (heavy) | Escherichia coli | 3 | 0.029 | 105 | matched |
|  |  |  | Klebsiella pneumoniae complex (heavy) | Not found | 0 | 0 |  | unmatched |
| 160 | 24MB125702 | polymicrobial | Escherichia coli (scanty) | Escherichia coli | 17 | 0.003 | 5999 | matched |
|  |  |  | Streptococcus salivarius group (heavy) | Streptococcus salivarius | 2811 | 0.469 |  | matched |
|  |  |  | Phocaeicola vulgatus (scanty) | Not found | 0 | 0 |  | unmatched |
| 161 | 24MB125897 | monomicrobial | Staphylococcus aureus (MRSA) (scanty) | Staphylococcus aureus | 54 | 1 | 54 | matched |
| 162 | 24MB126562 | monomicrobial | Staphylococcus aureus (MRSA) (moderate) | Staphylococcus aureus | 6114 | 1 | 6114 | matched |
| 163 | 24MB126914 | monomicrobial | Group G Streptococcus (heavy) | Streptococcus dysgalactiae | 5361 | 1 | 5361 | matched |
| 164 | 24MB127112 | polymicrobial | Escherichia coli (heavy) | Escherichia coli | 14 | 0.002 | 7914 | matched |
|  |  |  | Streptococcus anginosus (moderate) | Streptococcus anginosus | 29 | 0.004 |  | matched |
|  |  |  | Bacteroides species (heavy) | Bacteroides thetaiotaomicron | 32 | 0.004 |  | matched |
|  |  |  | Eggerthella lenta (heavy) | Not found | 0 | 0 |  | unmatched |
| 165 | 24MB127113 | polymicrobial | Streptococcus mitis group (heavy) | Streptococcus oralis | 523 | 0.093 | 5619 | matched |
|  |  |  | Enterococcus faecium (heavy) | Enterococcus faecium | 1661 | 0.296 |  | matched |
|  |  |  | Corynebacterium striatum (moderate) | Corynebacterium striatum | 62 | 0.011 |  | matched |
|  |  |  | Candida albicans (moderate) | Candida africana/ ablicans | 26 | 1 | 26 | matched |
| 166 | 24MB127767 | monomicrobial | Group G Streptococcus (heavy) | Streptococcus dysgalactiae | 10934 | 1 | 10934 | matched |
| 167 | 24MB127701 | monomicrobial | Klebsiella pneumoniae complex (heavy) | Klebsiella pneumoniae | 1381 | 0.486 | 2845 | matched |
| 168 | 24MB127956 | monomicrobial | Streptococcus constellatus (scanty) | Streptococcus constellatus | 16 | 0.005 | 2938 | matched |
| 169 | 24MB128107 | monomicrobial | Staphylococcus aureus (MRSA) (heavy) | Staphylococcus aureus | 5095 | 1 | 5095 | matched |
| 170 | 24MB128166 | polymicrobial | Salmonella species, Group D (heavy) | Salmonella enterica | 878 | 0.266 | 3300 | matched |
|  |  |  | Staphylococcus aureus (MRSA) (heavy) | Staphylococcus aureus | 2422 | 0.734 |  | matched |
| 171 | 24MB128227 | polymicrobial | Salmonella species, Group D (heavy) | Salmonella enterica | 166 | 0.251 | 662 | matched |
|  |  |  | Staphylococcus aureus (MRSA) (heavy) | Staphylococcus aureus | 496 | 0.749 |  | matched |
| 172 | 24MB128250 | monomicrobial | Parvimonas micra (scanty) | Parvimonas micra | 77 | 0.019 | 4037 | matched |
