## Supplemental Table 1. Primer sequences of ID and AMR primer set for "Optimizing and Evaluating Nanopore-Based Targeted and Metagenomic Sequencing Workflows for Rapid Diagnosis of Acute Invasive Infections from Normally Sterile Body Fluids"

| Primer set | Primer  name | Sequence (5' to 3') | Amplicon size (bp) | Targeted  AMR  genes | Working concentration (μM) | References |
| --- | --- | --- | --- | --- | --- | --- |
| ID primer set | 16S-27F-YM | AGAGTTTGATYMTGGCTCAG | 1500 | Bacterial 16S rRNA | 2 | 1 |
|  | 16S-1492R | ACGGYTACCTTGTTACGACTT |  |  |  |  |
|  | ITS1-27F | TACGTCCCTGCCCTTTGTAC | 200-500 | Fungal ITS | 1 | 2, 3 |
|  | ITS4 | TCCTCCGCTTATTGATATGC |  |  |  |  |
| AMR primer set | MecA-F | GGCTATCGTGTCACAATCGTT | 689 | *MecA* | 1 | 4 |
|  | MecA-R | TCACCTTGTCCGTAACCTGA |  |  |  |  |
|  | VanA-F | TCTGCAATAGAGATAGCCGC | 377 | *VanA* | 1 | 5 |
|  | VanA-R | GGAGTAGCTATCCCAGCATT |  |  |  |  |
|  | VanB-F | CATCGCCGTCCCCGAATTTCAAA | 297 | *VanB* | 1 | 6 |
|  | VanB-R | GATGCGGAAGATACCGTGGCT |  |  |  |  |
|  | CTX-MA1 | SCSATGTGCAGYACCAGTAA | 450 | *blaCTX-M* | 4 | 7 |
|  | CTX-MA2 | CCGCRATATGRTTGGTGGTG |  |  |  |  |
|  | CTX-M  Gp8/25-F | AACRCRCAGACGCTCTAC | 326 | *blaCTX-M-8, blaCTX-M-25, blaCTX-M-26 and blaCTX-M-39 to blaCTX-M-41* | 2 | 8 |
|  | CTX-M  Gp8/25-R | TCGAGCCGGAASGTGTYAT |  |  |  |  |
|  | TEM-F | CATTTCCGTGTCGCCCTTATTC | 800 | *blaTEM* | 1 | 9 |
|  | TEM-R | CGTTCATCCATAGTTGCCTGAC |  |  |  |  |
|  | SHV-F | AGCCGCTTGAGCAAATTAAAC | 713 | *blaSHV* | 1 | 9 |
|  | SHV-R | ATCCCGCAGATAAATCACCAC |  |  |  |  |
|  | OXA-F | GGCACCAGATTCAACTTTCAAG | 564 | *blaOXA-1* | 1 | 9 |
|  | OXA-R | GACCCCAAGTTTCCTGTAAGTG |  |  |  |  |
|  | KPC-F | TGTCACTGTATCGCCGTC | 900 | *blaKPC* | 1 | 10 |
|  | KPC-R | CTCAGTGCTCTACAGAAAACC |  |  |  |  |
|  | IMP-F | GAAGGCGTTTATGTTCATAC | 587 | *blaIMP* | 1 | 10 |
|  | IMP-R | GTACGTTTCAAGAGTGATGC |  |  |  |  |
|  | VIM-F | GTTTGGTCGCATATCGCAAC | 389 | *blaVIM* | 1 | 10 |
|  | VIM-R | AATGCGCAGCACCAGGATAG |  |  |  |  |
|  | NDM-F | GCAGCTTGTCGGCCATGCGGGC | 782 | *blaNDM* | 1 | 10 |
|  | NDM-R | GGTCGCGAAGCTGAGCACCGCAT |  |  |  |  |
|  | OXA-48-F | GCTTGATCGCCCTCGATT | 281 | *blaOXA-48* | 1 | 11 |
|  | OXA-48-R | GATTTGCTCCGTGGCCGAAA |  |  |  |  |
|  | ACC-F | AACAGCCTCAGCAGCCGGTTA | 346 | *blaACC* | 1 | 12 |
|  | ACC-R | TTCGCCGCAATCATCCCTAGC |  |  |  |  |
|  | FOX-F | GCCGAGGCTTACGGGATCAAG | 247 | *blaFOX-1 to 9* | 1 | 13 |
|  | FOX-R | CAAAGCGCGTAACCGGATTGG |  |  |  |  |
|  | MOX-F | GCAACAACGACAATCCATCCT | 895 | *blaMOX-1, blaMOX-2, blaCMY-1, blaCMY-8 to blaCMY-11 and blaCMY-19* | 1 | 8 |
|  | MOX-R | GGGATAGGCGTAACTCTCCCAA |  |  |  |  |
|  | DHA-F | AACTTTCACAGGTGTGCTGGGT | 405 | *blaDHA-1, blaDHA-2* | 1 | 12 |
|  | DHA-R | CCGTACGCATACTGGCTTTGC |  |  |  |  |
|  | CIT-F | CGAAGAGGCAATGACCAGAC | 538 | *blaLAT-1 to blaLAT-3, blaBIL-1, blaCMY-2 to blaCMY-7, blaCMY-12 to blaCMY-18 and blaCMY-21 to blaCMY-23* | 1 | 8 |
|  | CIT-R | ACGGACAGGGTTAGGATAGY |  |  |  |  |
|  | EBC-F | TCGGTAAAGCCGATGTTGCGG | 302 | *blaMIR-1, blaACT-1* | 1 | 12 |
|  | EBC-R | CTTCCACTGCGGCTGCCAGTT |  |  |  |  |
