## Supplemental 4: The concordance between NMgS and culture in pathogen identification for "Optimizing and Evaluating Nanopore-Based Targeted and Metagenomic Sequencing Workflows for Rapid Diagnosis of Acute Invasive Infections from Normally Sterile Body Fluids"

| **Supplemental 4: The concordance between NMgS and culture in pathogen identification** | | | | | |  |  |  |
| --- | --- | --- | --- | --- | --- | --- | --- | --- |
| **No.** | **Sample ID** | **No. of cultured species** | **culture** | **Kraken2 + Bracken (c=0.1, t=10)** | **Read count** | **Relative abundance** | **Total classified reads** | **concardance** |
| 1 | 23MB047690 | monomicrobial | Staphylococcus aureus (MRSA) (scanty) | Not found | 0 | 0 | 0 | unmatched |
| 2 | 23MB047738 | monomicrobial | Streptococcus agalactiae (Group B streptococcus) (heavy) | Streptococcus sp. | 1167 | 1 | 1167 | identified at genus level only |
| 3 | 23MB047824 | monomicrobial | Streptococcus pyogenes (Group A streptococcus) (moderate) | Streptococcus pyogenes | 22720 | 1 | 22720 | matched |
| 4 | 23MB047892 | monomicrobial | Staphylococcus aureus (heavy) | Staphylococcus aureus | 24822 | 1 | 24822 | matched |
| 5 | 23MB087252 | monomicrobial | Escherichia coli (scanty) | Not found | 0 | 0 | 0 | unmatched |
| 6 | 23MB087291 | monomicrobial | Candida albicans (Scanty) | Not found | 0 | 0 | 0 | unmatched |
| 7 | 23MB087709 | monomicrobial | Klebsiella pneumoniae complex (heavy) | Klebsiella pneumoniae | 22598 | 1 | 22598 | matched |
| 8 | 23MB087820 | monomicrobial | Klebsiella pneumoniae complex (scanty) | Not found | 0 | 0 | 0 | unmatched |
| 9 | 23MB087882 | monomicrobial | Staphylococcus aureus (scanty) | Staphylococcus aureus | 4147 | 1 | 4147 | matched |
| 10 | 23MB101355 | monomicrobial | Bacteroides fragilis (moderate) | Bacteroides fragilis | 1388 | 0.98 | 1410 | matched |
| 11 | 23MB101356 | monomicrobial | Staphylococcus aureus (heavy) | Staphylococcus aureus | 21876 | 1 | 21876 | matched |
| 12 | 23MB101447 | monomicrobial | Streptococcus dysgalactiae subsp. equisimilis (heavy) | Streptococcus dysgalactiae | 440 | 1 | 440 | matched |
| 13 | 23MB101616 | monomicrobial | Staphylococcus aureus (scanty) | Staphylococcus aureus | 164 | 0.72 | 227 | matched |
| 14 | 23MB110604 | monomicrobial | Campylobacter fetus (isolated) | Not found | 0 | 0 | 0 | unmatched |
| 15 | 23MB110726 | monomicrobial | Corynebacterium striatum (scanty) | Corynebacterium striatum | 119 | 1 | 119 | matched |
| 16 | 23M2010586 | polymicrobial | Enterococcus faecium (Heavy) | Enterococcus faecium | 15340 | 0.945 | 16232 | matched |
|  |  |  | Escherichia coli (Scanty) | Not found | 0 | 0 |  | unmatched |
|  |  |  | Klebsiella variicola (Scanty) | Not found | 0 | 0 |  | unmatched |
|  |  |  | Enterococcus raffinosus (Moderate) | Enterococcus raffinosus | 476 | 0.029 |  | matched |
|  |  |  | Clostridium perfringens (Scanty) | Clostridium perfringens | 42 | 0.003 |  | matched |
|  |  |  | Alpha-haemolytic Streptococci (Scanty) | Streptococcus sanguinis | 42 | 0.003 |  | matched |
|  |  |  | Proteus species (Scanty) | Not found | 0 | 0 |  | unmatched |
|  |  |  | Streptococcus infantarius (Scanty) | Streptococcus infantarius | 270 | 0.017 |  | matched |
| 17 | 23M2010695 | polymicrobial | Escherichia coli (Heavy) | Escherichia coli | 333 | 0.011 | 31376 | matched |
|  |  |  | Enterococcus faecalis (Moderate) | Enterococcus faecalis | 26402 | 0.841 |  | matched |
|  |  |  | Enterococcus avium (Scanty) | Enterococcus avium | 4258 | 0.136 |  | matched |
| 18 | 23M2010728 | polymicrobial | Escherichia coli (Moderate) | Escherichia coli | 25095 | 0.916 | 27385 | matched |
|  |  |  | Proteus mirabilis (Moderate) | Not found | 0 | 0 |  | unmatched |
|  |  |  | Klebsiella pneumoniae (Scanty) | Klebsiella sp. | 23 | 0.0008 |  | identified at genus level only |
|  |  |  | Clostridium perfringens (Scanty) | Clostridium perfringens | 75 | 0.003 |  | matched |
| 19 | 23M2011494 | polymicrobial | Escherichia coli (Scanty) | Not found | 0 | 0 | 99 | unmatched |
|  |  |  | Bacteroides fragilis (Scanty) | Bacteroides sp. | 17 | 0.172 |  | identified at genus level only |
|  |  |  | Coagulase negative Staphylococcus (Scanty) | Not found | 0 | 0 |  | unmatched |
|  |  |  | Streptococcus constellatus (Scanty) | Not found | 0 | 0 |  | unmatched |
| 20 | 23M2011587 | polymicrobial | Corynebacterium aurimucosum (Moderate) | Corynebacterium aurimucosum | 227 | 0.383 | 593 | matched |
|  |  |  | Coagulase negative Staphylococcus (Heavy) | Staphylococcus epidermidis | 366 | 0.617 |  | matched |
| 21 | 23M2102729 | polymicrobial | Escherichia coli (Heavy) | Escherichia coli | 1357 | 0.085 | 16023 | matched |
|  |  |  | Pseudomonas aeruginosa (Scanty) | Pseudomonas aeruginosa | 107 | 0.007 |  | matched |
|  |  |  | Streptococcus constellatus (Scanty) | Streptococcus anginosus group | 29 | 0.002 |  | identified at genus level only |
| 22 | 23M2102735 | monomicrobial | Candida albicans (Scanty) | Not found | 0 | 0 | 0 | unmatched |
| 23 | 23M2102736 | monomicrobial | Escherichia coli | Not found | 0 | 0 | 0 | unmatched |
| 24 | 23M2102738 | polymicrobial | Escherichia coli (Moderate) | Escherichia coli | 104 | 0.016 | 6672 | matched |
|  |  |  | Acinetobacter calcoaceticus-baumannii complex (Heavy) | Not found | 0 | 0 |  | unmatched |
|  |  |  | Klebsiella pneumoniae (Scanty) | Not found | 0 | 0 |  | unmatched |
|  |  |  | Lysinibacillus species (Scanty) | Not found | 0 | 0 |  | unmatched |
| 25 | 23M2102747 | monomicrobial | Coagulase negative Staphylococcus | Staphylococcus warneri | 67 | 0.313 | 214 | matched |
|  |  |  |  | Staphylococcus epidermidis | 22 | 0.103 |  |  |
| 26 | 23M2102754 | monomicrobial | Bacteroides pyogenes (Heavy) | Not found | 0 | 0 | 1663 | unmatched |
| 27 | 23M2102780 | monomicrobial | Enterococcus gallinarum | Not found | 0 | 0 | 0 | unmatched |
| 28 | 23M2102832 | monomicrobial | Staphylococcus aureus (MRSA) (Heavy) | Staphylococcus aureus | 5589 | 1 | 5589 | matched |
| 29 | 23M2102908 | monomicrobial | Bacteroides fragilis | Not found | 0 | 0 | 0 | unmatched |
| 30 | 23M2102914 | monomicrobial | Staphylococcus aureus (MRSA) | Staphylococcus aureus | 1087 | 0.845 | 1287 | matched |
| 31 | 23M2102931 | monomicrobial | Pseudomonas aeruginosa (Heavy) | Pseudomonas aeruginosa | 31739 | 0.997 | 31849 | matched |
| 32 | 23M4201725 | monomicrobial | Streptococcus intermedius (Heavy) | Streptococcus intermedius | 28 | 1 | 28 | matched |
| 33 | 23M9006507 | monomicrobial | Staphylococcus aureus (Moderate) | Not found | 0 | 0 | 0 | unmatched |
| 34 | 23M2104323 | polymicrobial | Escherichia coli (Heavy) | Not found | 0 | 0 | 70 | unmatched |
|  |  |  | Bacteroides fragilis (Heavy) | Bacteroides fragilis | 42 | 0.6 |  | matched |
|  |  |  | Enterococcus faecalis (Scanty) | Not found | 0 | 0 |  | unmatched |
| 35 | 23M2104297 | monomicrobial | Staphylococcus aureus | Not found | 0 | 0 | 0 | unmatched |
| 36 | 23M2104285 | monomicrobial | Acinetobacter calcoaceticus-baumannii complex (Moderate) | Acinetobacter pittii | 87 | 1 | 87 | matched |
| 37 | 23M2104292 | monomicrobial | Coagulase negative Staphylococcus | Not found | 0 | 0 | 0 | unmatched |
| 38 | 23M2104271 | monomicrobial | Streptococcus mitis group (Scanty) | Not found | 0 | 0 | 0 | unmatched |
| 39 | 23M2104267 | monomicrobial | Escherichia coli (Scanty) | Not found | 0 | 0 | 0 | unmatched |
| 40 | 23M2104266 | polymicrobial | Candida albicans (Scanty) | Not found | 0 | 0 | 0 | unmatched |
|  |  |  | Candida tropicalis (Scanty) | Not found | 0 | 0 |  | unmatched |
| 41 | 23M2104242 | monomicrobial | Escherichia coli (Scanty) | Not found | 0 | 0 | 0 | unmatched |
| 42 | 23M2104135 | monomicrobial | Candida parapsilosis | Not found | 0 | 0 | 0 | unmatched |
| 43 | 23M2104041 | monomicrobial | Coagulase negative Staphylococcus | Not found | 0 | 0 | 0 | unmatched |
| 44 | 23M2104033 | monomicrobial | Streptococcus anginosus | Not found | 0 | 0 | 0 | unmatched |
| 45 | 23M2103978 | monomicrobial | Streptococcus dysgalactiae (Scanty) | Streptococcus dysgalactiae | 128 | 1 | 128 | matched |
| 46 | 23M2103952 | monomicrobial | Corynebacterium striatum | Not found | 0 | 0 | 0 | unmatched |
| 47 | 23M2103944 | monomicrobial | Escherichia coli (Heavy) | Escherichia coli | 33196 | 0.99 | 33443 | matched |
| 48 | 23M2104372 | monomicrobial | Staphylococcus aureus (Scanty) | Not found | 0 | 0 | 0 | unmatched |
| 49 | 23M2104408 | monomicrobial | Streptococcus parasanguinis | Not found | 0 | 0 | 0 | unmatched |
| 50 | 23M2104413 | polymicrobial | Escherichia coli (Moderate) | Not found | 0 | 0 | 199 | unmatched |
|  |  |  | Peptostreptococcus anaerobius (Scanty) | Not found | 0 | 0 |  | unmatched |
|  |  |  | Actinomyces turicensis (Scanty) | Not found | 0 | 0 |  | unmatched |
|  |  |  | Prevotella intermedia (Moderate) | Prevotella intermedia | 73 | 0.367 |  | matched |
|  |  |  | Bacteroides cellulosilyticus (Moderate) | Not found | 0 | 0 |  | unmatched |
|  |  |  | Bacteroides ovatus (Scanty) | Not found | 0 | 0 |  | unmatched |
| 51 | 23M2104421 | monomicrobial | Staphylococcus lugdunensis (Scanty) | Not found | 0 | 0 | 0 | unmatched |
| 52 | 23M2104412 | polymicrobial | Streptococcus anginosus (Scanty) | Not found | 0 | 0 | 144 | unmatched |
|  |  |  | Rothia mucilaginosa (Scanty) | Rothia dentocariosa | 76 | 0.53 |  | matched |
|  |  |  | Streptococcus mitis group (Scanty) | Not found | 0 | 0 |  | unmatched |
|  |  |  | Staphylococcus aureus | Not found | 0 | 0 |  | unmatched |
|  |  |  | Streptococcus vestibularis | Not found | 0 | 0 |  | unmatched |
|  |  |  | Lactobacillus salivarius | Not found | 0 | 0 |  | unmatched |
| 53 | 24MB001633 | monomicrobial | Staphylococcus species (isolated) | Not found | 0 | 0 | 0 | unmatched |
| 54 | 24MB011005 | polymicrobial | Streptococcus agalactiae (Group B streptococcus) (scanty) | Not found | 0 | 0 | 0 | unmatched |
|  |  |  | Viridans streptococcus (heavy) | Not found | 0 | 0 |  | unmatched |
| 55 | 24MB011006 | monomicrobial | Staphylococcus aureus (MRSA) (scanty) | Not found | 0 | 0 | 0 | unmatched |
| 56 | 24MB011017 | monomicrobial | Klebsiella pneumoniae complex (scanty) | Not found | 0 | 0 | 0 | unmatched |
| 57 | 24MB011020 | monomicrobial | Enterococcus faecalis (moderate) | Enterococcus faecalis | 6539 | 1 | 6539 | matched |
| 58 | 24MB011073 | polymicrobial | Staphylococcus aureus (scanty) | Not found | 0 | 0 | 0 | unmatched |
|  |  |  | Streptococcus agalactiae (scanty) | Not found | 0 | 0 |  | unmatched |
| 59 | 24MB011062 | polymicrobial | Escherichia coli (scanty) | Not found | 0 | 0 | 0 | unmatched |
|  |  |  | Enterococcus faecium (isolated) | Not found | 0 | 0 |  | unmatched |
| 60 | 24MB011066 | monomicrobial | Pseudomonas aeruginosa (heavy) | Pseudomonas aeruginosa | 1790 | 1 | 1790 | matched |
| 61 | 24MB011104 | polymicrobial | Streptococcus anginosus group (heavy) | Streptococcus intermedius | 3302 | 0.074 | 44618 | matched |
|  |  |  | Parvimonas micra (heavy) | Parvimonas micra | 40234 | 0.902 |  | matched |
| 62 | 24MB011155 | polymicrobial | Escherichia coli (heavy) | Escherichia coli | 300 | 0.014 | 21228 | matched |
|  |  |  | Enterococcus faecium (heavy) | Enterococcus faecium | 20514 | 0.966 |  | matched |
|  |  |  | Acinetobacter baumannii complex (scanty) | Not found | 0 | 0 |  | unmatched |
|  |  |  | Burkholderia cepacia complex (scanty) | Not found | 0 | 0 |  | unmatched |
|  |  |  | Candida albicans (scanty) | Candida albicans | 207 | 0.01 |  | matched |
| 63 | 24MB011121 | polymicrobial | Klebsiella species (moderate) | Klebsiella sp. | 19 | 0.001 | 15776 | identified at genus level only |
|  |  |  | Escherichia coli (heavy) | Escherichia coli | 160 | 0.01 |  | matched |
|  |  |  | Enterococcus raffinosus (heavy) | Enterococcus raffinosus | 15428 | 0.978 |  | matched |
|  |  |  | Clostridium perfringes (scanty) | Not found | 0 | 0 |  | unmatched |
|  |  |  | Pseudomonas aeruginosa (moderate) | Not found | 0 | 0 |  | unmatched |
| 64 | 24MB011122 | monomicrobial | Escherichia coli (heavy) | Escherichia coli | 268 | 1 | 268 | matched |
| 65 | 24MB011188 | monomicrobial | Escherichia coli (heavy) | Not found | 0 | 0 | 0 | unmatched |
| 66 | 24MB011103 | monomicrobial | Escherichia coli (scanty) | Not found | 0 | 0 | 0 | unmatched |
| 67 | 24MB011255 | monomicrobial | Pseudomonas aeruginosa (scanty) | Not found | 0 | 0 | 0 | unmatched |
| 68 | 24MB011363 | polymicrobial | Escherichia coli (heavy) | Escherichia coli | 173 | 0.901 | 192 | matched |
|  |  |  | Proteus mirabilis (heavy) | Proteus mirabilis | 19 | 0.099 |  | matched |
|  |  |  | Candida tropicalis (scanty) | Not found | 0 | 0 |  | unmatched |
| 69 | 24MB011403 | polymicrobial | Escherichia coli (heavy) | Escherichia coli | 140 | 0.543 | 258 | matched |
|  |  |  | Enterococcus faecalis (heavy) | Enterococcus faecalis | 118 | 0.457 |  | matched |
|  |  |  | Klebsiella oxytoca (heavy) | Not found | 0 | 0 |  | unmatched |
| 70 | 24MB011364 | polymicrobial | Klebsiella pneumoniae complex (heavy) | Klebsiella sp. | 38 | 1 | 38 | identified at genus level only |
|  |  |  | Pseudomonas aeruginosa (heavy) | Not found | 0 | 0 |  | unmatched |
| 71 | 24MB011412 | monomicrobial | Escherichia coli (heavy) | Not found | 0 | 0 | 0 | unmatched |
| 72 | 24MB011541 | monomicrobial | Viridans streptococcus (heavy) | Streptococcus gordonii | 7108 | 0.992 | 7168 | matched |
| 73 | 24MB011567 | monomicrobial | Phocaeicola vulgatus (heavy) | Phocaeicola vulgatus | 18449 | 0.925 | 19949 | matched |
| 74 | 24MB011665 | monomicrobial | Klebsiella pneumoniae complex (moderate) | Klebsiella sp. | 24 | 0.96 | 25 | identified at genus level only |
| 75 | 24MB011666 | monomicrobial | Staphylococcus aureus (heavy) | Not found | 0 | 0 | 0 | unmatched |
| 76 | 24MB011667 | monomicrobial | Klebsiella pneumoniae complex (heavy) | Not found | 0 | 0 | 0 | unmatched |
| 77 | 24MB011662 | monomicrobial | Pseudomonas aeruginosa (scanty) | Not found | 0 | 0 | 0 | unmatched |
| 78 | 24MB011753 | monomicrobial | Klebsiella aerogenes (heavy) | Klebsiella aerogenes | 18331 | 0.977 | 18764 | matched |
| 79 | 24MB011679 | polymicrobial | Acinetobacter baumannii complex (scanty) | Not found | 0 | 0 | 421 | unmatched |
|  |  |  | Staphylococcus aureus (MRSA) (moderate) | Not found | 0 | 0 |  | unmatched |
|  |  |  | Enterococcus casseliflavus (heavy) | Enterococcus casseliflavus | 163 | 0.387 |  | matched |
|  |  |  | Lactobacillus paracasei (heavy) | Lacticaseibacillus paracasei | 109 | 0.259 |  | matched |
|  |  |  | Candida tropicalis (heavy) | Candida sp. | 38 | 0.09 |  | identified at genus level only |
| 80 | 24MB011678 | polymicrobial | Enterobacter species (heavy) | Enterobacter roggenkampii | 121 | 0.01 | 12529 | matched |
|  |  |  | Kluyvera georgiana (moderate) | Kluyvera sp. | 33 | 0.003 |  | identified at genus level only |
|  |  |  | GPC in chains (heavy) | Streptococcus pasteurianus | 11081 | 0.884 |  | matched |
|  |  |  |  | Streptococcus anginosus group | 1313 | 0.105 |  |  |
| 81 | 24MB011637 | polymicrobial | Pseudomonas aeruginosa (heavy) | Pseudomonas aeruginosa | 4361 | 0.186 | 23499 | matched |
|  |  |  | Escherichia coli (moderate) | Escherichia coli | 17901 | 0.762 |  | matched |
|  |  |  | Klebsiella pneumoniae complex (heavy) | Klebsiella quasipneumoniae | 28 | 0.001 |  | matched |
|  |  |  | Bacteroides species (heavy) | Bacteroides thetaiotaomicron | 36 | 0.002 |  | matched |
|  |  |  | Enterococcus faecium (scanty) | Enterococcus faecium | 32 | 0.001 |  | matched |
| 82 | 23MB170626 | monomicrobial | Pseudomonas aeruginosa (isolated) | Not found | 0 | 0 | 0 | unmatched |
| 83 | 23MB170293 | monomicrobial | Staphylococcus aureus (scanty) | Staphylococcus sp. | 40 | 0.506 | 79 | identified at genus level only |
| 84 | 23MB184156 | monomicrobial | Staphylococcus aureus (moderate) | Staphylococcus aureus | 2333 | 1 | 2333 | matched |
| 85 | 23MB184024 | monomicrobial | Staphylococcus aureus (MRSA) (moderate) | Not found | 0 | 0 | 0 | unmatched |
| 86 | 23MB170776 | polymicrobial | Pseudomonas aeruginosa (heavy) | Pseudomonas aeruginosa | 118 | 1 | 118 | matched |
|  |  |  | Klebsiella pneumoniae complex (isolated) | Not found | 0 | 0 |  | unmatched |
| 87 | 23MB170782 | monomicrobial | Staphylococcus aureus (MRSA) (scanty) | Not found | 0 | 0 | 0 | unmatched |
| 88 | 23MB170774 | monomicrobial | Streptococcus anginosus group (scanty) | Streptococcus intermedius | 15 | 0.5 | 30 | matched |
| 89 | 23MB170490 | polymicrobial | Bacteroides fragilis (moderate) | Bacteroides fragilis | 57 | 1 | 57 | matched |
|  |  |  | Escherichia coli (moderate) | Not found | 0 | 0 |  | unmatched |
| 90 | 23MB170331 | monomicrobial | Aeromonas species (heavy) | Aeromonas veronii | 109959 | 0.975 | 112775 | matched |
|  |  |  |  | Aeromonas caviae | 1036 | 0.009 |  |  |
| 91 | 23MB184255 | polymicrobial | Streptococcus anginosus group (heavy) | Streptococcus anginosus | 302 | 0.019 | 15983 | matched |
|  |  |  | Escherichia coli (heavy) | Not found | 0 | 0 |  | unmatched |
|  |  |  | Bacteroides thetaiotaomicron (heavy) | Bacteroides thetaiotaomicron | 47 | 0.003 |  | matched |
| 92 | 23MB184185 | monomicrobial | Streptococcus anginosus group (scanty) | Not found | 0 | 0 | 0 | unmatched |
| 93 | 24MB017342 | polymicrobial | Klebsiella pneumoniae complex (moderate) | Klebsiella sp. | 16 | 0.001 | 15640 | identified at genus level only |
|  |  |  | Escherichia coli (heavy) | Not found | 0 | 0 |  | unmatched |
|  |  |  | Enterococcus avium (heavy) | Enterococcus avium | 14995 | 0.959 |  | matched |
| 94 | 24MB017347 | polymicrobial | Escherichia coli (moderate) | Escherichia coli | 4596 | 0.131 | 35069 | matched |
|  |  |  | Aeromonas species (heavy) | Aeromonas caviae | 25195 | 0.718 |  | matched |
|  |  |  | Klebsiella oxytoca (heavy) | Klebsiella oxytoca | 724 | 0.021 |  | matched |
|  |  |  | Enterococcus species (scanty) | Enterococcus raffinosus | 62 | 0.002 |  | matched |
|  |  |  | Proteus species (scanty) | Proteus penneri | 350 | 0.01 |  | matched |
| 95 | 24MB017428 | monomicrobial | Klebsiella pneumoniae complex (scanty) | Not found | 0 | 0 | 0 | unmatched |
| 96 | 24MB017427 | monomicrobial | Escherichia coli (heavy) | Escherichia coli | 13856 | 1 | 13856 | matched |
| 97 | 24MB017494 | monomicrobial | Escherichia coli (moderate) | Not found | 0 | 0 | 0 | unmatched |
| 98 | 24MB017555 | polymicrobial | Escherichia coli (heavy) | Not found | 0 | 0 | 0 | unmatched |
|  |  |  | Streptococcus anginosus group (moderate) | Not found | 0 | 0 |  | unmatched |
|  |  |  | Bacteroides species (heavy) | Not found | 0 | 0 |  | unmatched |
| 99 | 24MB017777 | polymicrobial | Streptococcus anginosus group (scanty) | Not found | 0 | 0 | 0 | unmatched |
|  |  |  | Peptoniphilus species (scanty) | Not found | 0 | 0 |  | unmatched |
| 100 | 24M2101353 | polymicrobial | Streptococcus vestibularis | Not found | 0 | 0 | 0 | unmatched |
|  |  |  | Streptococcus salivarius | Not found | 0 | 0 |  | unmatched |
| 101 | 24M2101323 | monomicrobial | Coagulase negative Staphylococcus (scanty) | Not found | 0 | 0 | 0 | unmatched |
| 102 | 24M2004435 | monomicrobial | Escherichia coli (moderate) | Escherichia coli | 657 | 1 | 657 | matched |
| 103 | 24M2004419 | polymicrobial | Escherichia coli (moderate) | Escherichia coli | 41330 | 0.964 | 42874 | matched |
|  |  |  | Enterococcus hirae (moderate) | Enterococcus hirae | 13 | 0.0003 |  | matched |
|  |  |  | Clostridium perfringens (moderate) | Clostridium sp. | 10 | 0.0002 |  | identified at genus level only |
| 104 | 24MB032054 | monomicrobial | Pseudomonas species | Pseudomonas species | 5768 | 0.995 | 5797 | matched |
| 105 | 24MB032068 | polymicrobial | Acinetobacter baumannii complex | Acinetobacter sp. | 18 | 0.305 | 59 | identified at genus level only |
|  |  |  | candida tropicalis | Not found | 0 | 0 |  | unmatched |
| 106 | 24MB032078 | monomicrobial | Viridans Streptococcus | Not found | 0 | 0 | 0 | unmatched |
| 107 | 24MB032085 | monomicrobial | Pseudomonas species (isolated) | Not found | 0 | 0 | 0 | unmatched |
| 108 | 24MB032086 | polymicrobial | Escherichia coli (heavy) | Not found | 0 | 0 | 1083 | unmatched |
|  |  |  | Pseudomonas aeruginosa (moderate) | Pseudomonas sp. | 80 | 0.074 |  | identified at genus level only |
|  |  |  | Streptococcus anginosus group (heavy) | Not found | 0 | 0 |  | unmatched |
|  |  |  | Bacteroides thetaiotaomicron (heavy) | Bacteroides thetaiotaomicron | 15 | 0.014 |  | matched |
| 109 | 24MB110434 | monomicrobial | Klebsiella pneumoniae complex (scanty) | Klebsiella sp. | 10 | 1 | 10 | identified at genus level only |
| 110 | 24MB110571 | monomicrobial | Staphylococcus aureus (heavy) | Not found | 0 | 0 | 0 | unmatched |
| 111 | 24MB110656 | monomicrobial | Staphylococcus aureus (moderate) | Not found | 0 | 0 | 0 | unmatched |
| 112 | 24MB110723 | monomicrobial | Citrobacter koseri (scanty) | Citrobacter sp. | 13 | 0.123 | 106 | identified at genus level only |
| 113 | 24MB111063 | monomicrobial | Enterococcus faecalis (moderate) | Not found | 0 | 0 | 0 | unmatched |
| 114 | 24MB111194 | polymicrobial | Pseudomonas aeruginosa (moderate) | Pseudomonas aeruginosa | 8662 | 0.304 | 28509 | matched |
|  |  |  | Morganella morganii (moderate) | Morganella morganii | 19509 | 0.684 |  | matched |
| 115 | 24MB111565 | polymicrobial | Morganella morganii (moderate) | Morganella morganii | 216 | 0.927 | 233 | matched |
|  |  |  | Escherichia coli (scanty) | Not found | 0 | 0 |  | unmatched |
| 116 | 24MB111308 | monomicrobial | Staphylococcus aureus (moderate) | Not found | 0 | 0 | 0 | unmatched |
| 117 | 24MB111311 | monomicrobial | Citrobacter freundii complex (moderate) | Not found | 0 | 0 | 0 | unmatched |
| 118 | 24MB111315 | polymicrobial | Escherichia coli (moderate) | Not found | 0 | 0 | 19485 | unmatched |
|  |  |  | Streptococcus constellatus (scanty) | Not found | 0 | 0 |  | unmatched |
|  |  |  | Parvimonas micra (heavy) | Parvimonas micra | 13770 | 0.707 |  | matched |
| 119 | 24MB111480 | monomicrobial | Staphylococcus aureus (moderate) | Staphylococcus aureus | 303 | 1 | 303 | matched |
| 120 | 24MB111876 | monomicrobial | Klebsiella pneumoniae complex (heavy) | Klebsiella sp. | 19 | 1 | 19 | identified at genus level only |
| 121 | 24MB111852 | monomicrobial | Klebsiella pneumoniae complex (moderate) | Klebsiella sp. | 64 | 1 | 64 | identified at genus level only |
| 122 | 24MB112057 | monomicrobial | Citrobacter koseri (scanty) | Citrobacter koseri | 1987 | 1 | 1987 | matched |
| 123 | 24MB112341 | polymicrobial | Acinetobacter calcoaceticus baumannii complex (scanty) | Not found | 0 | 0 | 0 | unmatched |
|  |  |  | Staphylococcus aureus (MRSA) (scanty) | Not found | 0 | 0 |  | unmatched |
|  |  |  | Bacteroides species (scanty) | Not found | 0 | 0 |  | unmatched |
| 124 | 24MB112432 | monomicrobial | Pseudomonas aeruginosa (scanty) | Pseudomonas aeruginosa | 93 | 1 | 93 | matched |
| 125 | 24MB112433 | monomicrobial | Pseudomonas aeruginosa (scanty) | Pseudomonas sp. | 138 | 1 | 138 | identified at genus level only |
| 126 | 24MB112478 | monomicrobial | Escherichia coli (scanty) | Not found | 0 | 0 | 0 | unmatched |
| 127 | 24MB111695 | monomicrobial | Campylobacter species (scanty) | Campylobacter fetus | 81 | 1 | 81 | matched |
| 128 | 24MB111720 | monomicrobial | Streptococcus anginosus (moderate) | Streptococcus anginosus | 128 | 1 | 128 | matched |
| 129 | 24MB113274 | monomicrobial | Klebsiella pneumoniae complex (moderate) | Klebsiella sp. | 76 | 1 | 76 | identified at genus level only |
| 130 | 24MB112605 | monomicrobial | Klebsiella pneumoniae complex (scanty) | Not found | 0 | 0 | 0 | unmatched |
| 131 | 24MB112927 | monomicrobial | Streptococcus milleri group (scanty) | Streptococcus intermedius | 64 | 0.753 | 85 | matched |
| 132 | 24MB113276 | polymicrobial | Morganella morganii (heavy) | Morganella morganii | 1337 | 0.04 | 33676 | matched |
|  |  |  | Streptococcus constellatus (scanty) | Not found | 0 | 0 |  | unmatched |
|  |  |  | Fusobacterium mortiferum (scanty) | Fusobacterium mortiferum | 96 | 0.003 |  | matched |
|  |  |  | Clostridium ramosum (Thomasclavelia ramosa) (scanty) | Thomasclavelia ramosa | 36 | 0.001 |  | matched |
| 133 | 24MB113346 | monomicrobial | Escherichia coli (scanty) | Not found | 0 | 0 | 0 | unmatched |
| 134 | 24MB113395 | monomicrobial | Klebsiella pneumoniae complex (moderate) | Klebsiella variicola | 136 | 1 | 136 | matched |
| 135 | 24MB113404 | monomicrobial | Streptococcus intermedius (moderate) | Streptococcus intermedius | 600 | 1 | 600 | matched |
| 136 | 24MB113415 | monomicrobial | Streptococcus agalactiae (scanty) | Streptococcus sp. | 10 | 1 | 10 | identified at genus level only |
| 137 | 24MB113419 | polymicrobial | Escherichia coli (scanty) | Not found | 0 | 0 | 0 | unmatched |
|  |  |  | Klebsiella pneumoniae complex (scanty) | Not found | 0 | 0 |  | unmatched |
| 138 | 24MB113466 | monomicrobial | Klebsiella pneumoniae complex (scanty) | Klebsiella sp. | 48 | 1 | 48 | identified at genus level only |
| 139 | 24MB117350 | monomicrobial | Streptococcus constellatus (heavy) | Not found | 0 | 0 | 0 | unmatched |
| 140 | 24MB117532 | monomicrobial | Salmonella species, Group D (moderate) | Not found | 0 | 0 | 0 | unmatched |
| 141 | 24MB117763 | monomicrobial | Klebsiella pneumoniae complex (scanty) | Not found | 0 | 0 | 0 | unmatched |
| 142 | 24MB117992 | polymicrobial | Escherichia coli (moderate) | Not found | 0 | 0 | 211 | unmatched |
|  |  |  | Enterococcus raffinosus (heavy) | Not found | 0 | 0 |  | unmatched |
| 143 | 24MB118151 | polymicrobial | Escherichia coli (moderate) | Escherichia coli | 644 | 0.068 | 9410 | matched |
|  |  |  | Klebsiella oxytoca (moderate) | Not found | 0 | 0 |  | unmatched |
|  |  |  | Enterococcus avium (scanty) | Enterococcus avium | 21 | 0.002 |  | matched |
| 144 | 24MB118533 | polymicrobial | Streptococcus anginosus (heavy) | Streptococcus anginosus | 72 | 0.212 | 340 | matched |
|  |  |  | Raoutella ornithinolytica (heavy) | Raoultella sp. | 152 | 0.447 |  | identified at genus level only |
|  |  |  | Bacteroides fragilis (scanty) | Bacteroides fragilis | 69 | 0.203 |  | matched |
| 145 | 24MB119459 | monomicrobial | Klebsiella pneumoniae complex (heavy) | Not found | 0 | 0 | 0 | unmatched |
| 146 | 24MB119528 | polymicrobial | Escherichia coli (heavy) | Not found | 0 | 0 | 182 | unmatched |
|  |  |  | Streptococcus anginosus (scanty) | Not found | 0 | 0 |  | unmatched |
|  |  |  | Bacteriodes caccae (heavy) | Not found | 0 | 0 |  | unmatched |
|  |  |  | Clostridium perfringens (scanty) | Not found | 0 | 0 |  | unmatched |
| 147 | 24MB120055 | monomicrobial | Escherichia coli (scanty) | Not found | 0 | 0 | 0 | unmatched |
| 148 | 24MB120463 | monomicrobial | Streptococcus agalactiae (scanty) | Not found | 0 | 0 | 0 | unmatched |
| 149 | 24MB120515 | polymicrobial | Escherichia coli (moderate) | Not found | 0 | 0 | 81128 | unmatched |
|  |  |  | Enterococcus avium (scanty) | Not found | 0 | 0 |  | unmatched |
|  |  |  | Clostridium innocuum (scanty) | Not found | 0 | 0 |  | unmatched |
|  |  |  | Peptostreptococcus anaerobius (scanty) | Not found | 0 | 0 |  | unmatched |
|  |  |  | Bacteriodes fragilis (heavy) | Bacteroides fragilis | 79364 | 0.978 |  | matched |
| 150 | 24MB120576 | monomicrobial | Staphylococcus aureus (moderate) | Staphylococcus sp. | 12 | 1 | 12 | identified at genus level only |
| 151 | 24MB122216 | monomicrobial | Staphylococcus aureus (MRSA) (heavy) | Staphylococcus aureus | 19502 | 1 | 19502 | matched |
| 152 | 24MB122452 | polymicrobial | Neisseria flavescens subflava group (moderate) | Not found | 0 | 0 | 0 | unmatched |
|  |  |  | Streptococcus mitis group (moderate) | Not found | 0 | 0 |  | unmatched |
|  |  |  | Candida glabrata (heavy) | Not found | 0 | 0 |  | unmatched |
| 153 | 24MB122527 | polymicrobial | Escherichia coli (moderate) | Not found | 0 | 0 | 0 | unmatched |
|  |  |  | Citrobacter koseri (moderate) | Not found | 0 | 0 |  | unmatched |
|  |  |  | Enterococcus raffinosus (moderate) | Not found | 0 | 0 |  | unmatched |
|  |  |  | Candida glabrata (heavy) | Not found | 0 | 0 |  | unmatched |
| 154 | 24MB122856 | monomicrobial | Salmonella species, Group B (moderate) | Salmonella sp. | 58 | 1 | 58 | identified at genus level only |
| 155 | 24MB122862 | monomicrobial | Klebsiella pneumoniae complex (scanty) | Klebsiella sp. | 77 | 1 | 77 | identified at genus level only |
| 156 | 24MB123950 | monomicrobial | Streptococcus intermedius (moderate) | Streptococcus intermedius | 359 | 1 | 359 | matched |
| 157 | 24MB123988 | monomicrobial | Pseudomonas aeruginosa (scanty) | Pseudomonas aeruginosa | 58 | 1 | 58 | matched |
| 158 | 24MB124304 | monomicrobial | Streptococcus intermedius (moderate) | Streptococcus intermedius | 257 | 1 | 257 | matched |
| 159 | 24MB125173 | polymicrobial | Escherichia coli (heavy) | Not found | 0 | 0 | 21113 | unmatched |
|  |  |  | Klebsiella pneumoniae complex (heavy) | Not found | 0 | 0 |  | unmatched |
| 160 | 24MB125702 | polymicrobial | Escherichia coli (scanty) | Not found | 0 | 0 | 20749 | unmatched |
|  |  |  | Streptococcus salivarius group (heavy) | Streptococcus salivarius | 1618 | 0.078 |  | matched |
|  |  |  | Phocaeicola vulgatus (scanty) | Not found | 0 | 0 |  | unmatched |
| 161 | 24MB125897 | monomicrobial | Staphylococcus aureus (MRSA) (scanty) | Not found | 0 | 0 | 0 | unmatched |
| 162 | 24MB126562 | monomicrobial | Staphylococcus aureus (MRSA) (moderate) | Staphylococcus aureus | 532 | 1 | 532 | matched |
| 163 | 24MB126914 | monomicrobial | Group G Streptococcus (heavy) | Streptococcus dysgalactiae | 39502 | 0.995 | 39705 | matched |
| 164 | 24MB127112 | polymicrobial | Escherichia coli (heavy) | Not found | 0 | 0 | 12692 | unmatched |
|  |  |  | Streptococcus anginosus (moderate) | Streptococcus anginosus | 91 | 0.007 |  | matched |
|  |  |  | Bacteroides species (heavy) | Bacteroides thetaiotaomicron | 170 | 0.013 |  | matched |
|  |  |  | Eggerthella lenta (heavy) | Eggerthella lenta | 14 | 0.001 |  | matched |
| 165 | 24MB127113 | polymicrobial | Streptococcus mitis group (heavy) | Streptococcus mitis | 179 | 0.028 | 6332 | matched |
|  |  |  | Enterococcus faecium (heavy) | Enterococcus faecium | 1773 | 0.28 |  | matched |
|  |  |  | Corynebacterium striatum (moderate) | Corynebacterium striatum | 190 | 0.03 |  | matched |
|  |  |  | Candida albicans (moderate) | Not found | 0 | 0 |  | unmatched |
| 166 | 24MB127767 | monomicrobial | Group G Streptococcus (heavy) | Streptococcus dysgalactiae | 6179 | 1 | 6179 | matched |
| 167 | 24MB127701 | monomicrobial | Klebsiella pneumoniae complex (heavy) | Klebsiella sp. | 36 | 0.571 | 63 | identified at genus level only |
| 168 | 24MB127956 | monomicrobial | Streptococcus constellatus (scanty) | Streptococcus constellatus | 705 | 0.016 | 42972 | matched |
| 169 | 24MB128107 | monomicrobial | Staphylococcus aureus (MRSA) (heavy) | Staphylococcus aureus | 37 | 1 | 37 | matched |
| 170 | 24MB128166 | polymicrobial | Salmonella species, Group D (heavy) | Salmonella sp. | 1811 | 0.506 | 3581 | identified at genus level only |
|  |  |  | Staphylococcus aureus (MRSA) (heavy) | Staphylococcus aureus | 1770 | 0.494 |  | matched |
| 171 | 24MB128227 | polymicrobial | Salmonella species, Group D (heavy) | Salmonella sp. | 1361 | 0.441 | 3086 | identified at genus level only |
|  |  |  | Staphylococcus aureus (MRSA) (heavy) | Staphylococcus aureus | 1725 | 0.559 |  | matched |
| 172 | 24MB128250 | monomicrobial | Parvimonas micra (scanty) | Parvimonas micra | 53 | 0.039 | 1346 | matched |
| 173 | 24MB128272 | polymicrobial | Escherichia coli (moderate) | Not found | 0 | 0 | 556 | unmatched |
|  |  |  | Pseudomonas aeruginosa (moderate) | Not found | 0 | 0 |  | unmatched |
|  |  |  | Streptococcus constellatus (scanty) | Not found | 0 | 0 |  | unmatched |
|  |  |  | Bacteroides uniformis (scanty) | Not found | 0 | 0 |  | unmatched |
| 174 | 24MB128288 | polymicrobial | Enterococcus faecium (scanty) | Not found | 0 | 0 | 71 | unmatched |
|  |  |  | Candida glabrata (scanty) | Not found | 0 | 0 |  | unmatched |
| 175 | 24MB128557 | polymicrobial | Staphylococcus aureus (MRSA) (heavy) | Not found | 0 | 0 | 0 | unmatched |
|  |  |  | Salmonella species, Group D (moderate) | Not found | 0 | 0 |  | unmatched |
| 176 | 24MB128570 | polymicrobial | Escherichia coli (heavy) | Not found | 0 | 0 | 13090 | unmatched |
|  |  |  | Streptococcus milleri group (heavy) | Not found | 0 | 0 |  | unmatched |
|  |  |  | Clostridium tertium (scanty) | Not found | 0 | 0 |  | unmatched |
| 177 | 24MB128773 | polymicrobial | Candida glabrata (heavy) | Nakaseomyces glabratus | 264 | 0.01 | 27537 | matched |
|  |  |  | Candida albicans (scanty) | Not found | 0 | 0 |  | unmatched |
