## Supplemental 1. Detailed protocol for NTS workflow for "Optimizing and Evaluating Nanopore-Based Targeted and Metagenomic Sequencing Workflows for Rapid Diagnosis of Acute Invasive Infections from Normally Sterile Body Fluids"

Multiplex PCR

DNA extraction was performed using the QIAamp BiOstic Bacteremia DNA Kit. Multiplex PCR was performed using the ID primer set for bacterial and fungal identification and the AMR primer set for antimicrobial resistance detection (primer sequences in Supplemental Table 1). Nanopore adapter sequences (5’-TTTCTGTTGGTGCTGATATTGC-3’ forward; 5’-ACTTGCCTGTCGCTCTATCTTC-3’ reverse) were appended to primers for subsequent barcoding PCR. ID and AMR PCR reactions were prepared separately, each containing 12.5 μl LongAmp Hot Start Taq 2× Master Mix, 2.5 μl primer mix, and 10 μl DNA. Both reactions amplified under the following conditions: 95°C for 3 min; 20 cycles of 95°C for 30 s, 55°C for 30 s, 65°C for 1 min 40 s; final extension at 65°C for 5 min. Two PCR products of each sample were pooled, purified with 35 μl of AMPure XP beads, and eluted in 24 μl nuclease-free water.

Library preparation and sequencing

Library preparation was conducted using the ligation sequencing kit (SQK-LSK110 for R9.4.1 or SQK-LSK114 for R10.4.1 flow cells) and the PCR Barcoding Expansion 1–96 kit (EXP-PBC096), with minor modifications. Barcoding PCR was performed by mixing 24 μl of purified multiplex PCR product with 25 μl of LongAmp Taq 2× Master Mix and 1 μl of a PCR barcode. Thermal cycling conditions were: 95°C for 3 min; 15 cycles of 95°C for 15 s, 62°C for 15 s, and 65°C for 2 min; followed by a final extension at 65°C for 5 min. Amplicons were purified using 40 μl of AMPure XP beads and eluted in 11 μl of nuclease-free water. After quantification and normalization, up to 24 barcoded samples were pooled, end-repaired, adapter-ligated, and sequenced on a GridION for 4 hours. During sequencing, “barcode both ends,” and “trim barcodes” options were enabled.
