## Supplemental 2. Detailed protocols for NMgS workflow for "Optimizing and Evaluating Nanopore-Based Targeted and Metagenomic Sequencing Workflows for Rapid Diagnosis of Acute Invasive Infections from Normally Sterile Body Fluids"

Host DNA depletion and DNA extraction

Host DNA depletion was performed before DNA extraction. For blood samples, 200 μl of hetasep was added to 1 ml of whole blood samples to remove red blood cells. After mixing, samples were centrifuged at 100 × g for 3 minutes and incubated at 37°C for 5 minutes. The supernatants were collected and centrifuged at 10,000 × g at 4°C for 5 minutes. For other body fluid types, 1 ml of the fluid was directly centrifuged at 10,000 × g at 4°C for 5 minutes. Pellets were resuspended in 1 ml of nuclease-free water and incubated for 5 minutes to induce osmotic shock on human cells, followed by centrifugation at 10,000 × g at 4°C for 5 minutes. Then, the resulting pellets were resuspended in 95 μl of 0.0125% saponin and incubated at room temperature for 5 minutes to further lyse the human cells. To crosslink host DNA, 5 μl of 0.4 mM PMAxx (20 μM) was added, followed by a 10-minute dark incubation and 15-minute light exposure on ice within 20 cm of a light source. Samples were washed with 1 ml saline, and DNA was extracted using the QIAamp BiOstic Bacteremia DNA Kit.

Library preparation and sequencing

Similarly, library preparation of NMgS workflow was also performed using ligation sequencing kit (SQK-LSK110 or SQK-LSK114) and PCR Barcoding Expansion 1-96 (EXP-PBC096) with modifications. Genomic DNA (26 μl) was fragmented and A-tailed by adding 7 μl of NEBNext Ultra II FS Reaction Buffer and 2 μl of FS Enzyme Mix, incubated at 37°C for 30 s and 65°C for 30 min. Samples were purified with 35 μl of AMPure XP beads and eluted in 15 μl of nuclease-free water. Barcode adaptor ligation was performed by adding 10 μl of barcode adapter and 25 μl of Blunt/TA Ligase Master Mix, incubated at room temperature for 10 min, followed by purification with 25 μl of AMPure XP beads and elution in 24 μl of nuclease-free water. Barcode PCR was performed as described in NTS workflow, but with 35 cycles. Amplicons were purified with 40 μl of AMPure XP beads and eluted in 11 μl of nuclease-free water.

Sample input for pooling was calculated as 1000 ng divided by the number of samples. The required volume per sample was determined by dividing the input mass by the DNA concentration. For end-repair, 1 μl of DNA Control Sample, 7 μl of Ultra II End-Prep Reaction Buffer, and 3 μl of End-Prep Enzyme Mix were added to 49 μl of pooled library (scaled proportionally if volume exceeded 49 μl), followed by incubation at 20°C for 5 min and 65°C for 5 min. The library was purified with 1× AMPure XP beads and eluted in 60 μl of nuclease-free water. Adapter ligation was performed by adding 5 μl of adapter mix, 25 μl of ligation buffer, and 10 μl of NEBNext Quick T4 DNA Ligase to 60 μl of library. After a 10-minute incubation at room temperature, the library was purified with 40 μl of AMPure XP beads, washed twice with 250 μl of short fragment buffer, and eluted in 13 μl of elution buffer. Sequencing was conducted on a GridION platform using the super-accurate (SUP) model for 4 hours. Adaptive sampling was enabled to deplete host DNA by targeting the human reference genome GRCh38.p13.

Sequencing data analysis

The sequencing reads were analyzed using an in-house analysis pipeline. Briefly, the sequencing reads were first filtered using NanoFilt v2.8.0 to remove reads with length below 200bp, followed by removal of human reads using Kraken 2 v2.1.3 with the human genome (GRCh38.p13) database. Then, the filtered reads were classified using Kraken 2 and with PlusPF database. The species abundance was re-estimated from the Kraken 2 results using Bracken v2.9. For AMR genes identification, the filtered reads were classified using blast+ with the NCBI Reference Gene Catalog, as described in the NTS workflow.
